# Drowning in the Lake Victoria basin: A systematic review of prevalence, risk factors, and interventions in East Africa

**DOI:** 10.1101/2025.06.29.25330530

**Authors:** Madeline Sellinger, Grace Guo, Kyra Guy, Frederick Oporia, Heather Wipfli

## Abstract

What is already known on this topic - *summarise the state of scientific knowledge on this subject before you did your study and why this study needed to be done*.

There is a significant body of research on drowning in low-middle-income countries with general knowledge of the demographic and behavioral risk factors and an emerging body of research on this topic specific to Uganda. This systematic review aims to gather the existing research for the Lake Victoria basin and highlight critical gaps in order to direct future research, policy, and interventions in the field.

What this study adds - *summarise what we now know because of this study that we did not know before*

Crucial gaps in content identified for the Lake Victoria basin included: studies with geographic focus in Kenya or Tanzania, studies of local populations, studies of direct drowning prevention interventions, studies on behavioral (non-demographic) risk factors such as alcohol use, and studies assessing community knowledge, attitudes, and beliefs related to drowning prevention and water safety.

How might this study affect research, practice, or policy? *Summarise the implications of this study*.

This study represents the first systematic review including both peer-reviewed and gray literature centered on the Lake Victoria basin, and indicates key gaps in knowledge that can direct future research efforts.

**Background:** With the highest number of drowning fatalities per square kilometer, Lake Victoria, the world’s second-largest body of fresh water, represents the most dangerous stretch of water. Despite the high burden of drowning in the region, data on drowning incidence and risk vary. This study aims to assess existing research on drowning risk, policy, and interventions in the Lake Victoria basin to identify key gaps and guide future research and policy recommendations.

**Methods:** We conducted a cumulative systematic review following Templier and Pare’s 2015 framework to collect all literature on drowning in Kenya, Uganda, and Tanzania. PubMed, Web of Science, Cochrane, Google Scholar, and EMBASE were searched from 1/1/1999 through 6/2/2025. Data on geographic scope and setting, community perspectives, and literature type were extracted, and articles were analyzed for content to determine emerging themes.

**Results:** We identified 43 studies, including 37 peer-reviewed articles. There were eight global studies, seven multi-country, twenty in Uganda, three in Kenya, and five in Tanzania. 17.9% (n=5) of articles studied local populations, 67.9% (n=19) regional, and 14.3% (n= 4) national populations. 30.2% (n=13) mentioned community knowledge or attitudes. Over half of the studies focused on prevalence (23.2%, n=10) or risk factors (41.8%, n=18), and 13.9% (n=6) on direct interventions.

**Conclusion:** Understanding existing research on drowning in the Lake Victoria basin is crucial to identifying gaps and planning future research and interventions. Strengthening surveillance systems and evaluating culturally relevant prevention strategies are critical next steps to address this neglected public health issue.

## Background

Drowning is the third leading cause of death from unintentional injury worldwide [1]. Globally, there are about 300,000 drowning deaths each year with about 92% of these occurring in low- and middle-income countries (LMICs) [52]. Drowning mortality rates in the World Health Organization’s (WHO) African region are estimated to be among the highest in the world, with 5.6 per 100,000 people dying from a drowning-related incident each year [52]. Lake Victoria, the world’s second-largest body of fresh water with a shoreline stretching across Kenya, Uganda, and Tanzania, represents the most dangerous stretch of water in the world in terms of fatalities per square kilometer, with estimated drowning mortality of 2.9, 4.4, and 3.9 per 100,000, respectively [2, 52]. These numbers are believed to be an underestimate given that they do not include drowning incidents from transportation or flood disasters, which are frequent and increasing across many LMICs [1]. Men made up an 81%, 85%, and 88% share of drowning deaths in Kenya, Uganda, and Tanzania respectively [52]. In Kenya, 25-34 year olds made up the largest share of drowning deaths at 18% [52]. In Uganda, ages 15-29 made up the largest share, at 38% [52]. Lastly, Tanzania’s data indicated that 21% of drowning deaths were children under 5, with those older making up the remaining 79% of drowning deaths [52]. Survey data from several LMICs that account for submersions resulting from floods and water transport incidents suggests a drowning mortality rate four or five times higher than the WHO’s estimates [3]. Furthermore, many LMICs lack proper surveillance and thus lack accurate data on injuries, especially in the WHO African region.

Most drowning-related research that has been conducted in sub-Saharan Africa has primarily focused on South Africa, and data from non-South African countries is generally collected from broader injury or mortality studies, without a specific focus on drowning [4, 50]. With the prominent role that Lake Victoria plays in the economy, transportation, and day-to-day activities for lakeside communities in Kenya, Uganda, and Tanzania, further analysis into prevalence, at-risk groups, and policy will enable targeted and impactful interventions in the region. The lack of surveillance data on drowning incidence has contributed to a neglect of drowning prevention research, with a paucity of research to assess drowning risk factors or intervention programs in the areas in which drowning is most prevalent.

The high burden of drowning in the Lake Victoria basin necessitates consistent research to inform the scale-up of drowning prevention and control programs and policies. To help prioritize future research, policy, and intervention for drowning prevention and control around the Lake Victoria basin, this cumulative systematic review was conducted following Templier and Pare’s 2015 framework to collect all published peer-reviewed data and gray literature around the topic of drowning in the countries comprising the Lake Victoria basin—Kenya, Uganda, and Tanzania.

## Methods

### Search Strategy and Selection Criteria

Using the keywords and phrases listed in Table 1, we searched PubMed, Web of Science Cochrane, and EMBASE (Table 1). Web of Science implicitly uses an “and” operator. PubMed applies an “and” operator between concepts - concepts are identified via automatic term mapping. In Cochrane, if two or more terms are entered, search will combine the terms with “AND” to find articles or selected fields where both terms appear. EMBASE does not use implicit Boolean operators. Search terms were entered as listed as separate searches. Literature found via Google Scholar was hand-searched. A separate search for each of the search terms was run in Google Scholar. Given the number of results generated, literature was hand-searched and selected based on inclusion and exclusion criteria, forgoing the Excel sorting process conducted for the other databases. Drowning prevention and control was defined as any measure, direct or indirect, by which drowning mortality can be reduced. Both unintentional and intentional drowning were included in the study. Search was conducted on March 21 2024, with update search conducted on June 2 2025. We reviewed all literature published from January 1, 1999, through June 2, 2025, including peer-reviewed journal articles and governmental, non-governmental, and academic reports and fact sheets. To ensure the capture of all information, we cross-referenced the bibliographies of reviewed articles. The search included English-language articles. No foreign language articles were identified. Literature meeting relevancy and eligibility, shown in Figure 1, were extrapolated into the literature database for analysis (Figure 1). The identification, screening, eligibility assessment, and inclusion of studies were conducted in accordance with the Preferred Reporting Items for Systematic Reviews and Meta-Analyses (PRISMA) guidelines. The review has been registered in INPLASY database: https://doi.org/10.37766/inplasy2025.6.0080 [55].

**Figure 1:**
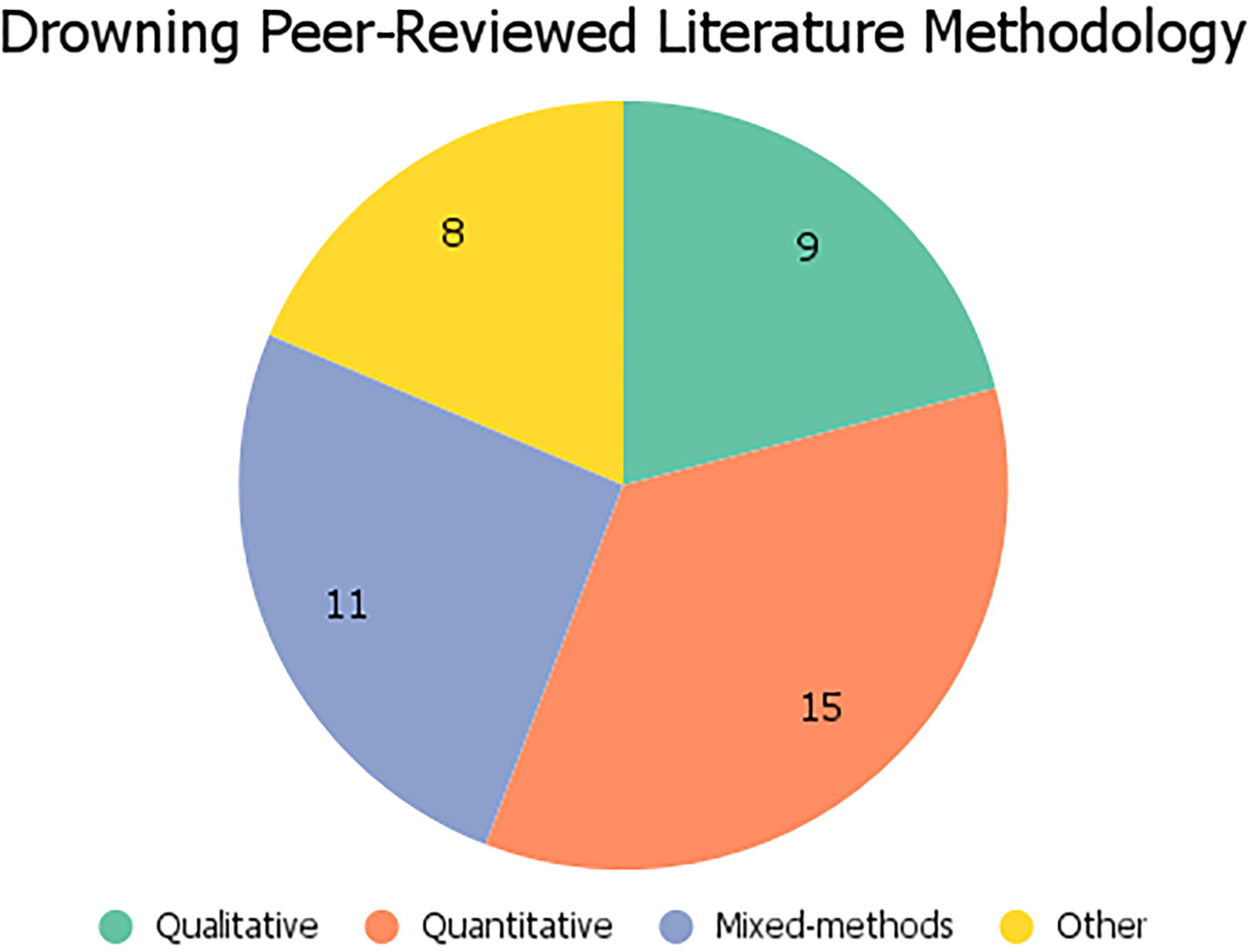
Systematic literature review following the PRISMA 2020 flow diagram [53].

**Table 1.**
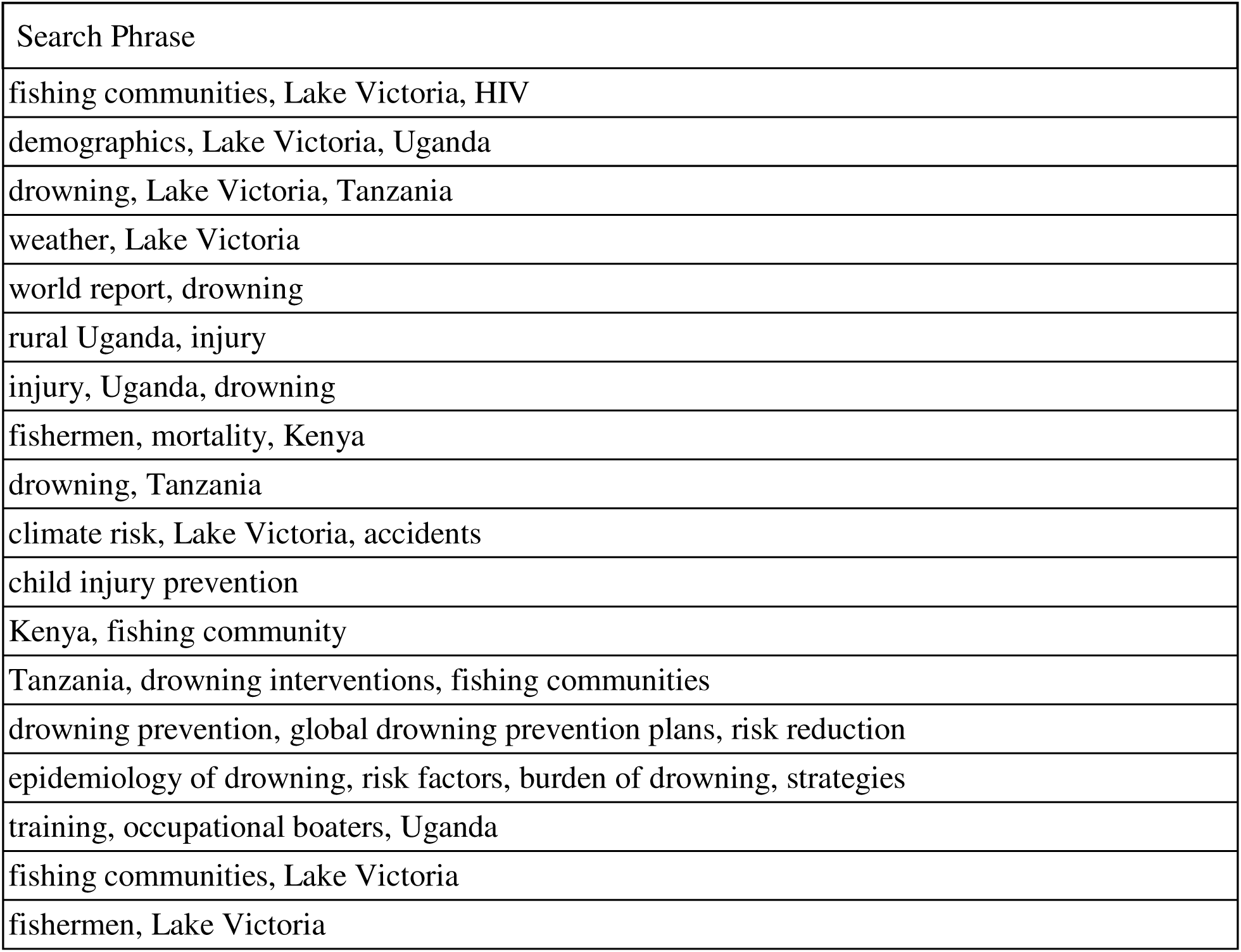
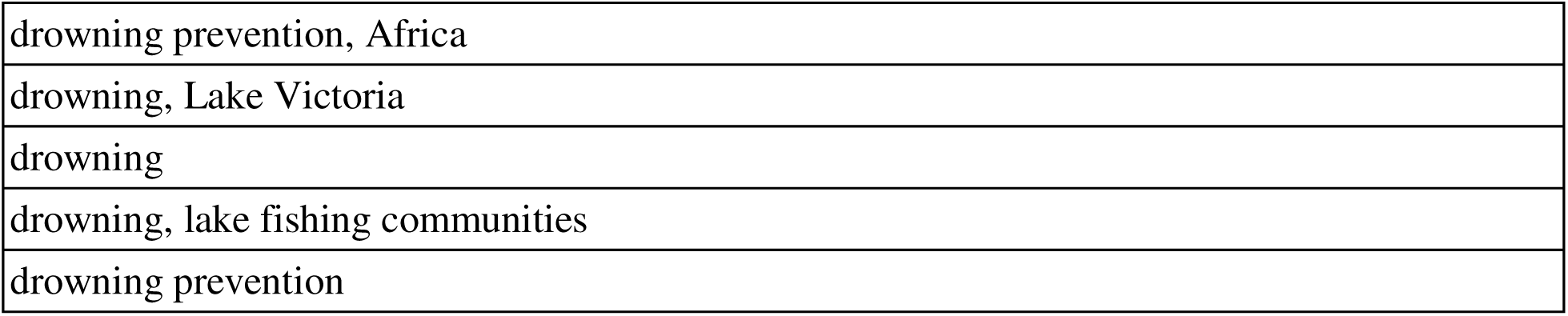
Search terms.

### Study Selection

One reviewer screened all articles based on title and abstract, and two reviewers screened 199 articles for full-text review using Excel, classifying each article as “Add to Database”, “Supplemental” - for general information that could be used in the manuscript writing process-“Irrelevant,” “Duplicated,” or “No Full Text Available” (MS & GG). Duplicates were identified using conditional formatting within Excel to identify articles with the same title, author, and DOI. Articles labeled as “Irrelevant,” “Duplicated,” or “No Full Text Available” in the screening process did not proceed to final analysis. Discrepancies were discussed and resolved through group consensus.

### Data Extraction and Quality Assessment

Inclusion criteria for this systematic review were the following: (1) any peer-reviewed or gray literature with a geographic setting in Uganda, Kenya, or Tanzania evaluating risk factors, prevalence, or policy for drowning, water safety, and fishing; (2) global studies of drowning risk factors, prevalence, or policy with specific discussion of Uganda, Kenya, or Tanzania; and (3) no restriction on race, gender, publication language, or date. Exclusion criteria are as follows: (1) study of drowning exclusively in countries outside the Lake Victoria basin; (2) articles without full text available; and (3) case reports. Features of the data collected were extracted from each study into a database spreadsheet in Excel. The following variables were collected as part of this process: study database, journal, year published, country setting, geographic scope within a country (if applicable), literature type (peer-reviewed or gray literature), inclusion of qualitative data on community knowledge or attitudes, and if drowning was the key focus of the article.

Geographic scope was defined as, for articles focusing on just one country, whether the study is “local” - one city/town-, regional-several cities/towns-, or national-multiple regions. In addition, each article was tagged as “prevalence,” “risk factors,” or “policy” to describe the primary focus of the data found in the article. Using the data collected, analysis was done to assess how much research has been published related to prevalence, risk factors, or policy, as well as the amount of data for a certain country and where that data is sourced geographically. Given the heterogeneity in study designs, a descriptive synthesis was conducted across all included studies, stratified by literature type (quantitative, qualitative, and mixed-methods)(Figure 4). For qualitative articles, thematic analysis was applied to extract key domains related to community knowledge, attitudes, and intervention priorities. Mixed Methods Appraisal Tool (MMAT) 2018 was used as a quality assessment tool (Table 2).

**Table 2.**
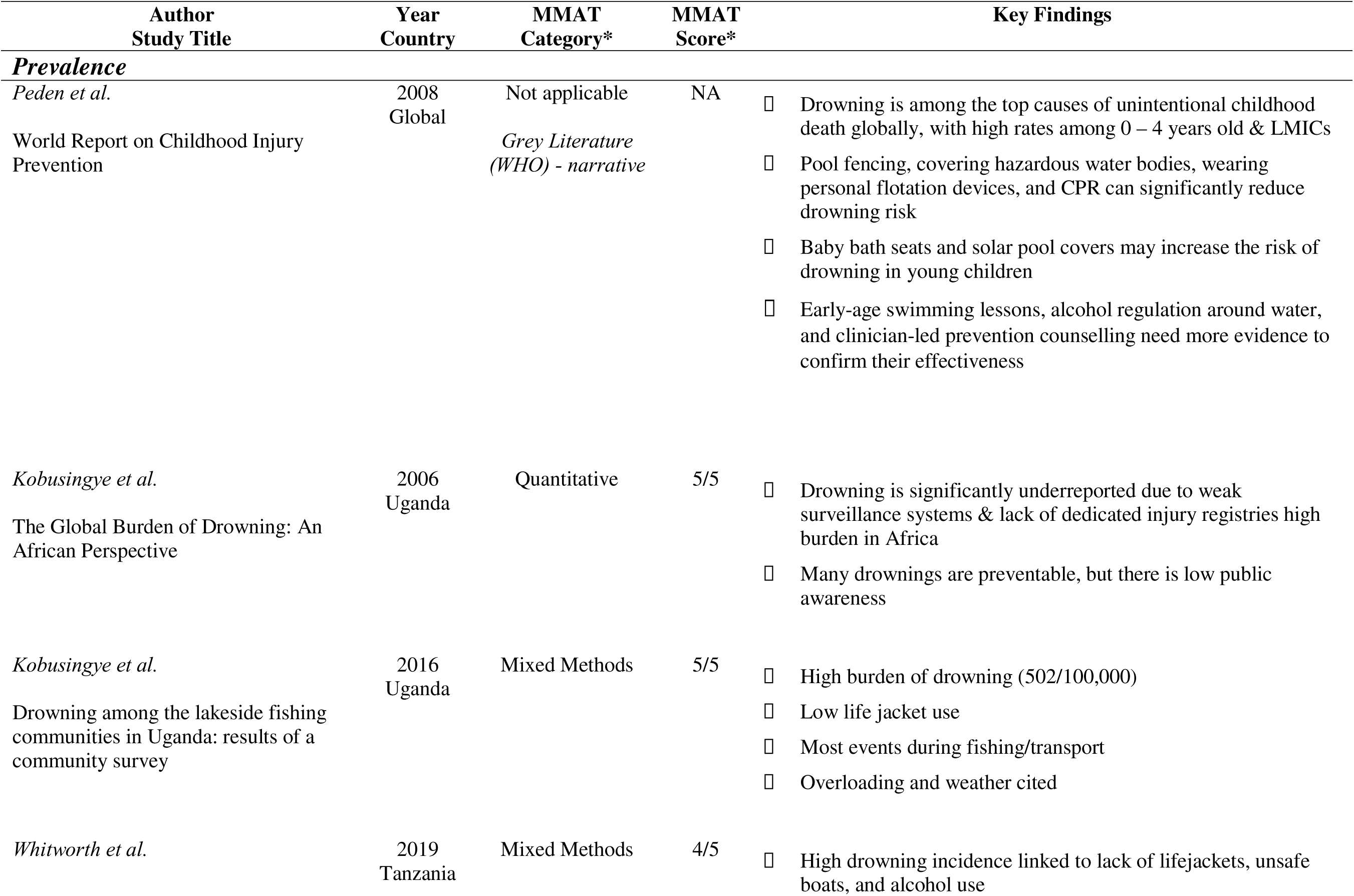
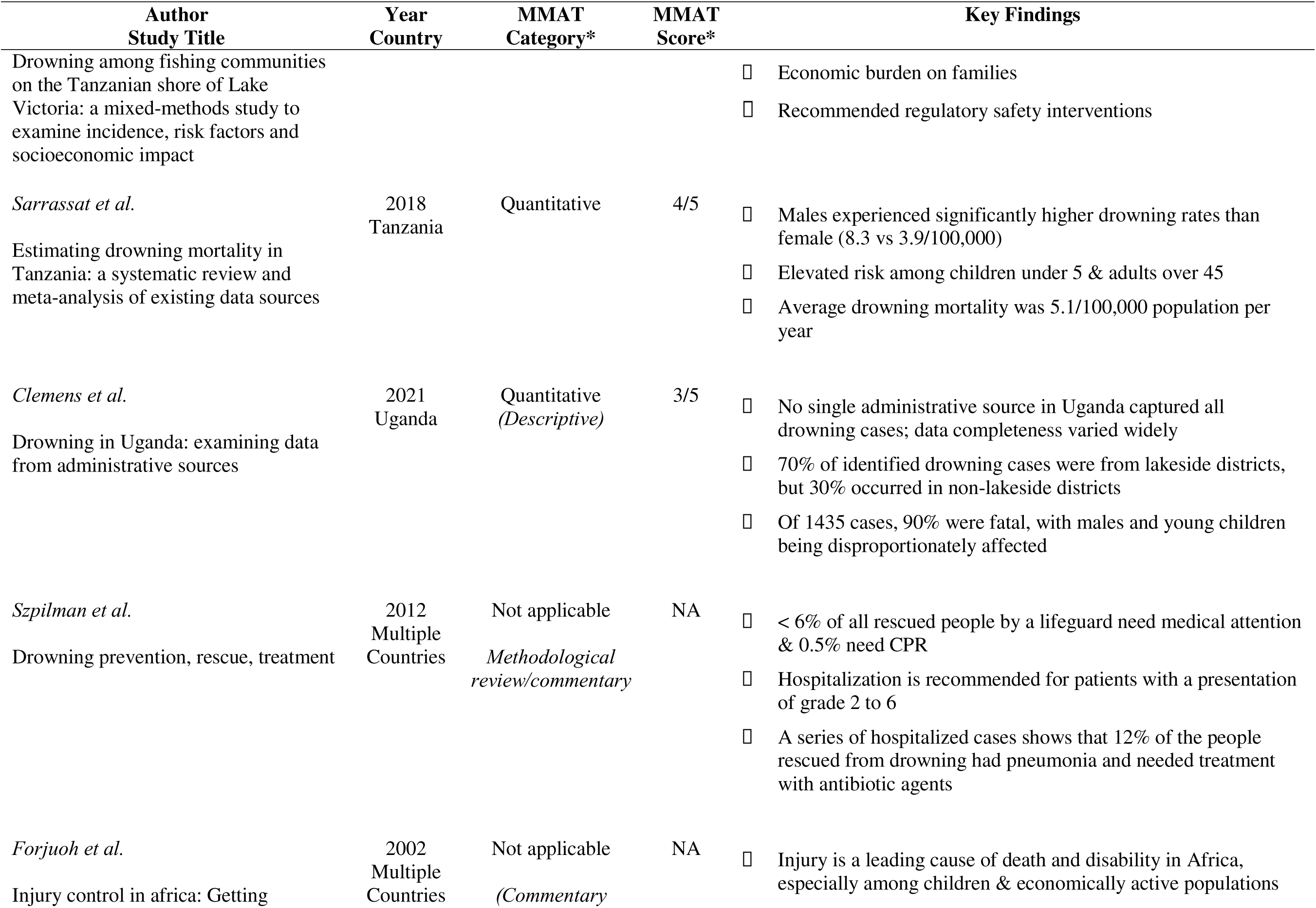
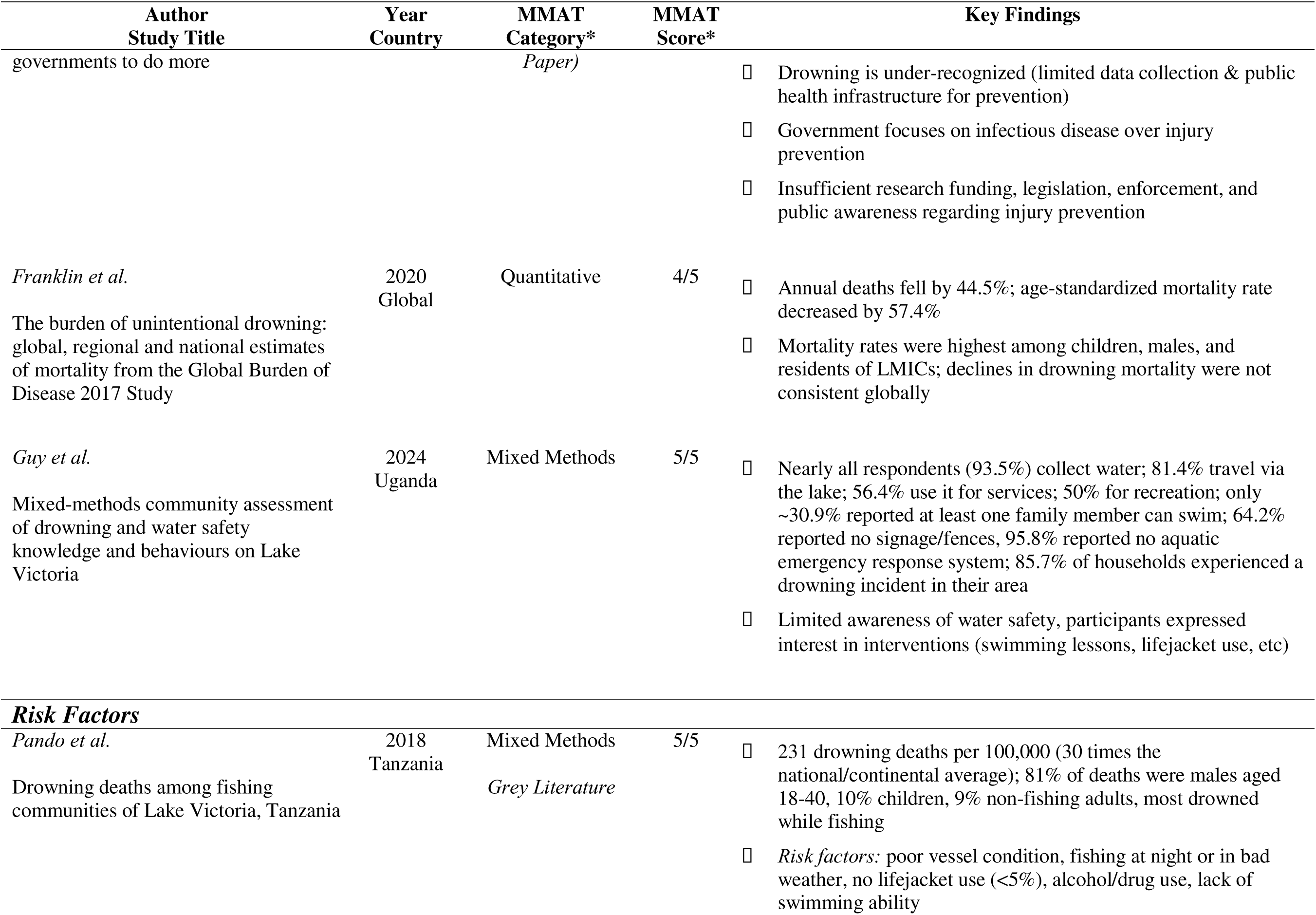
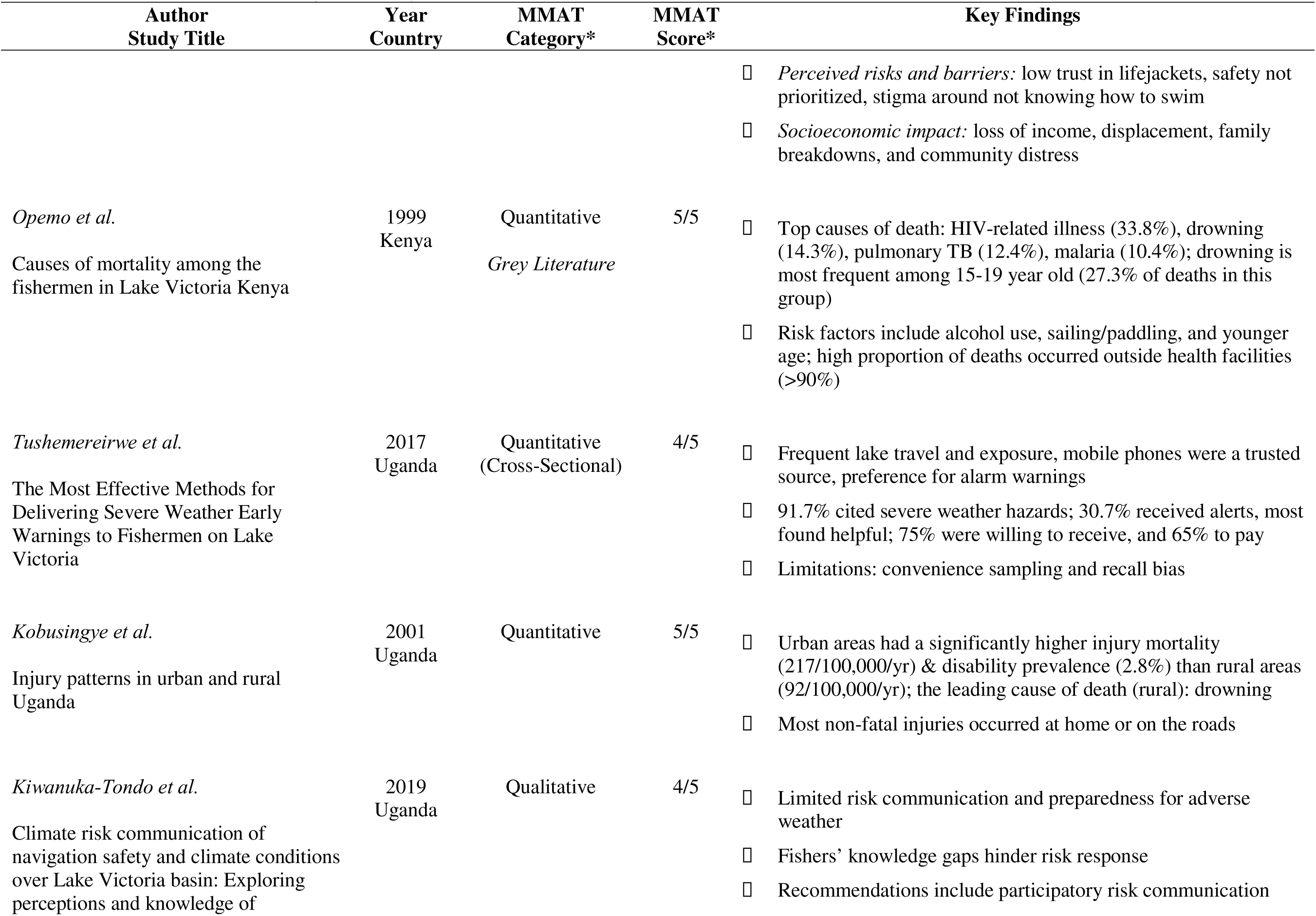
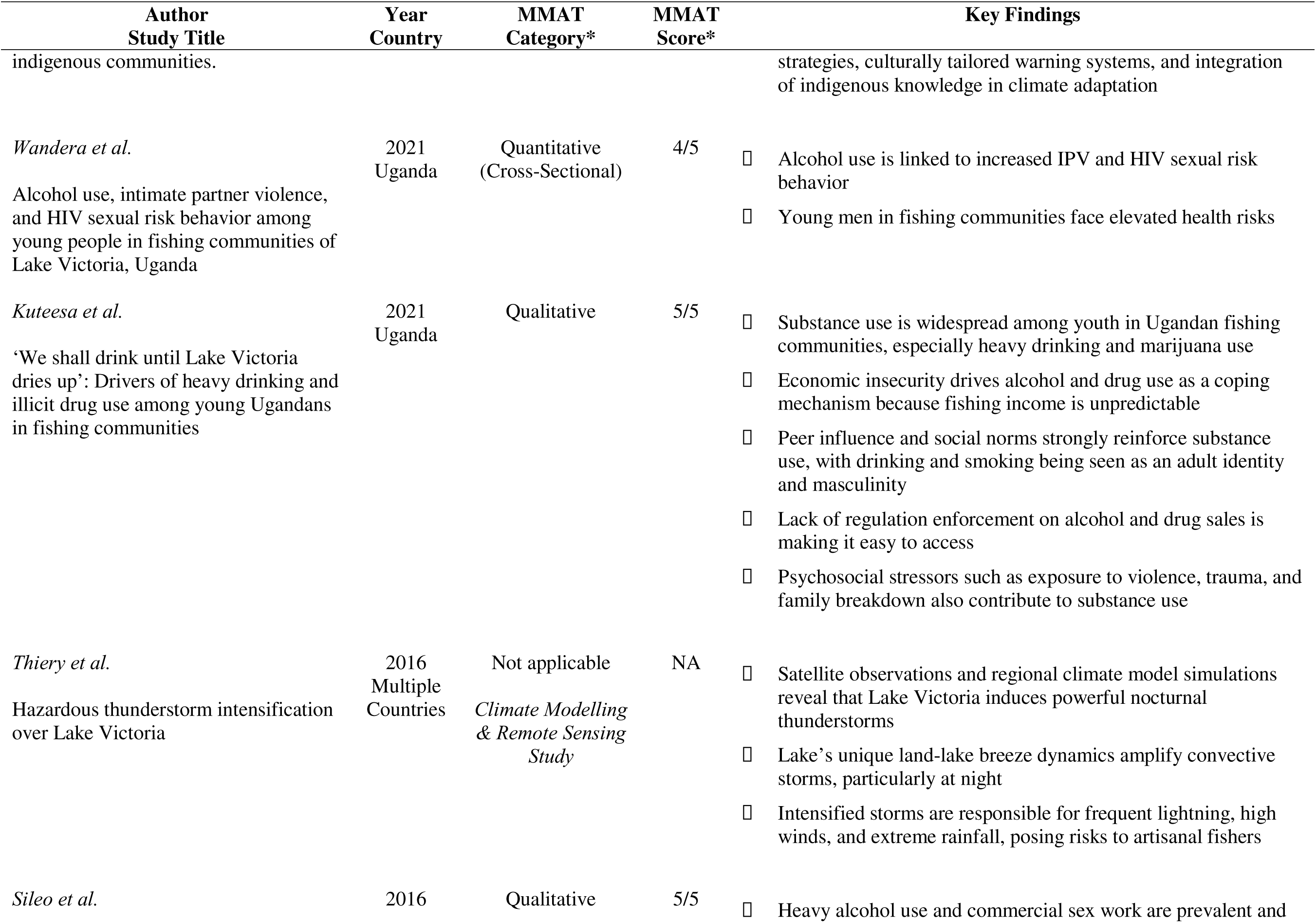
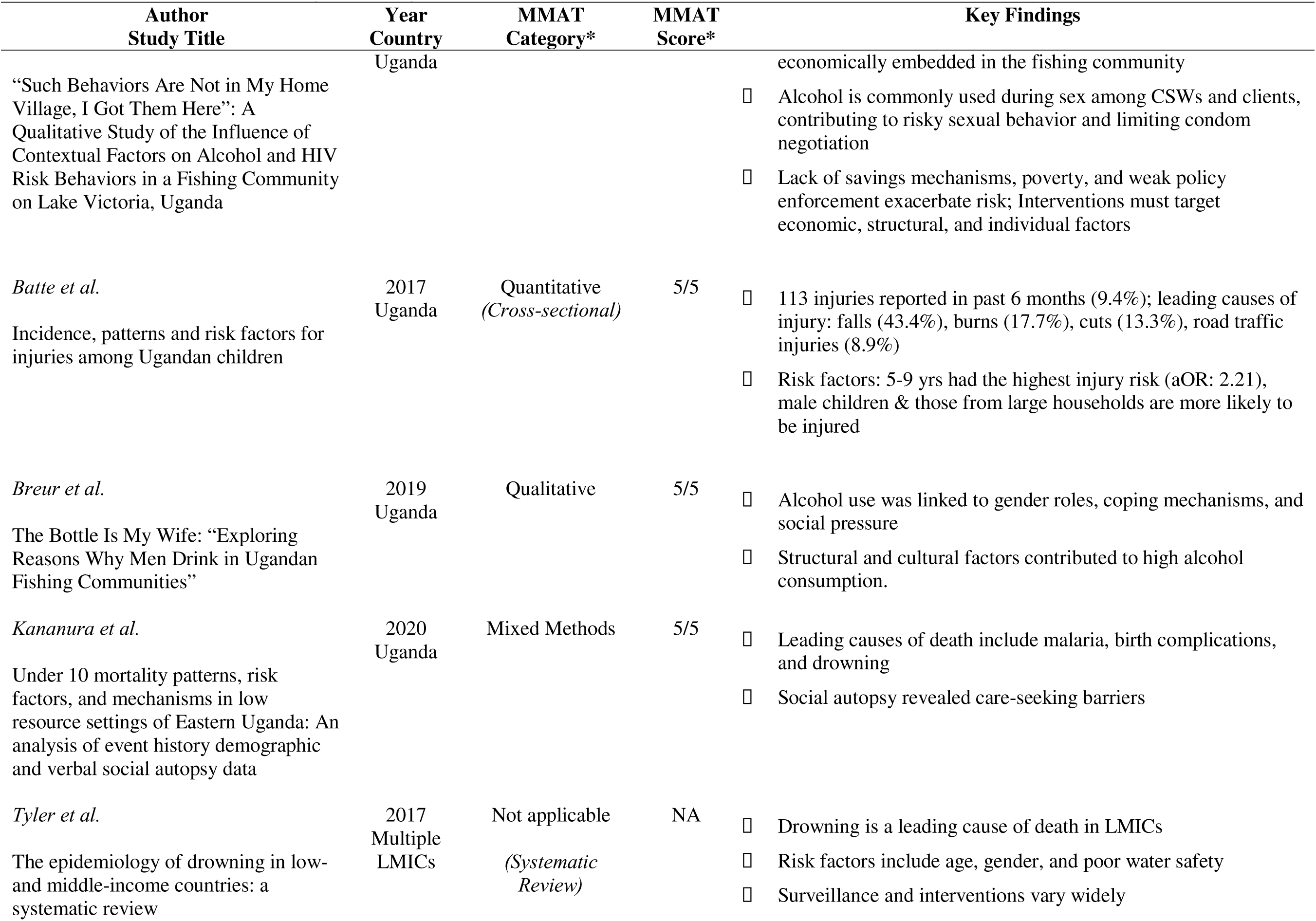
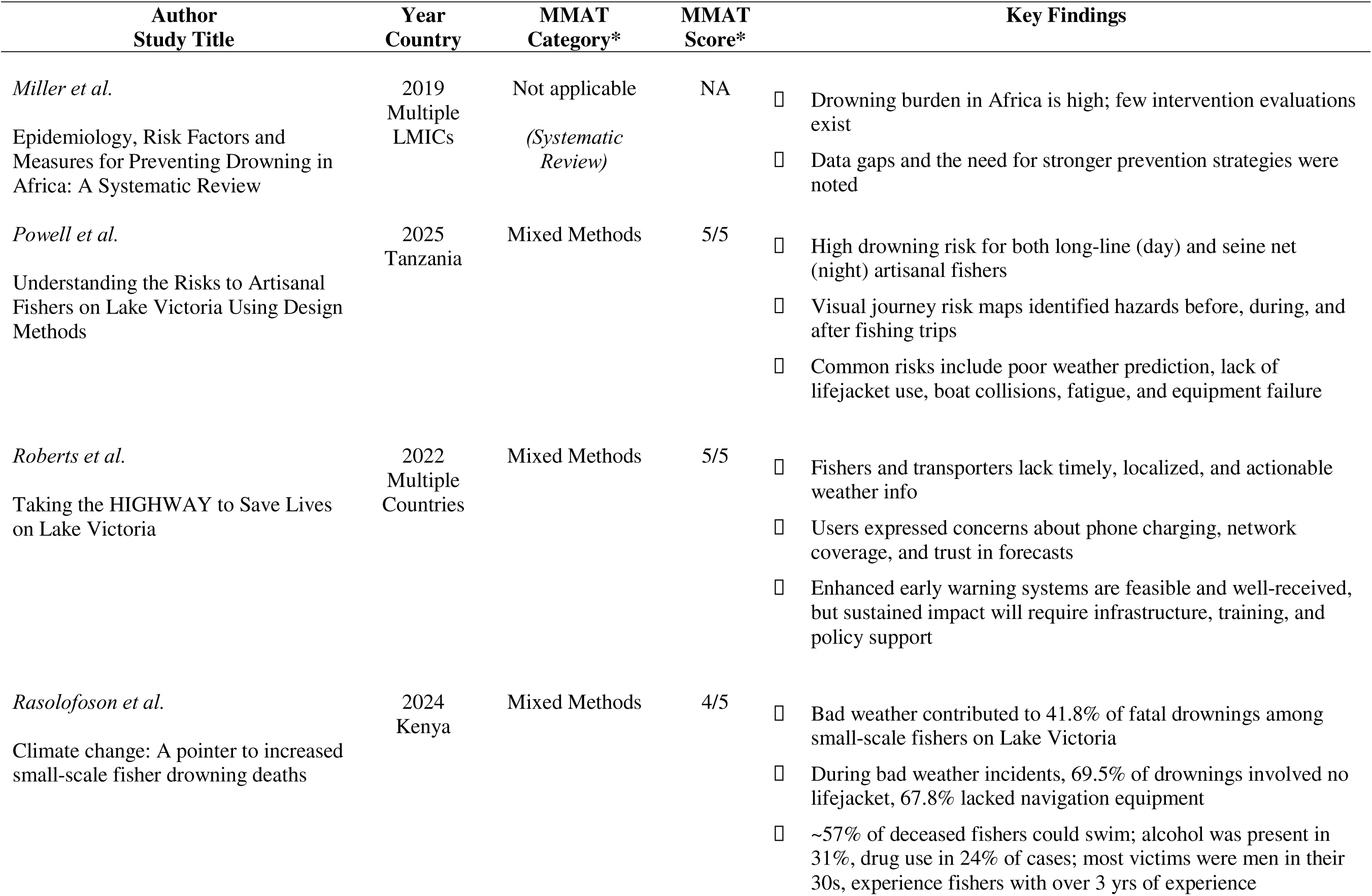
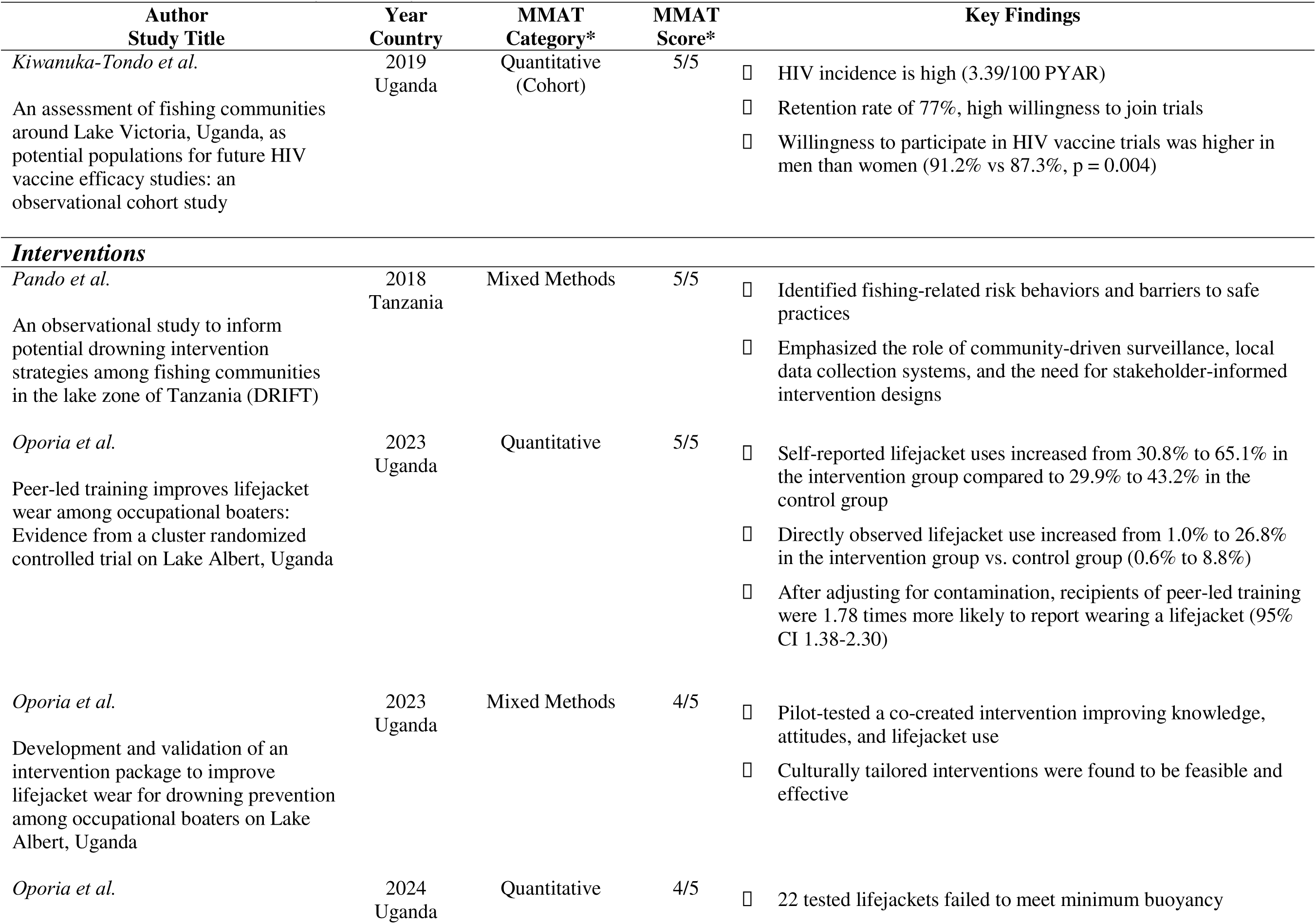
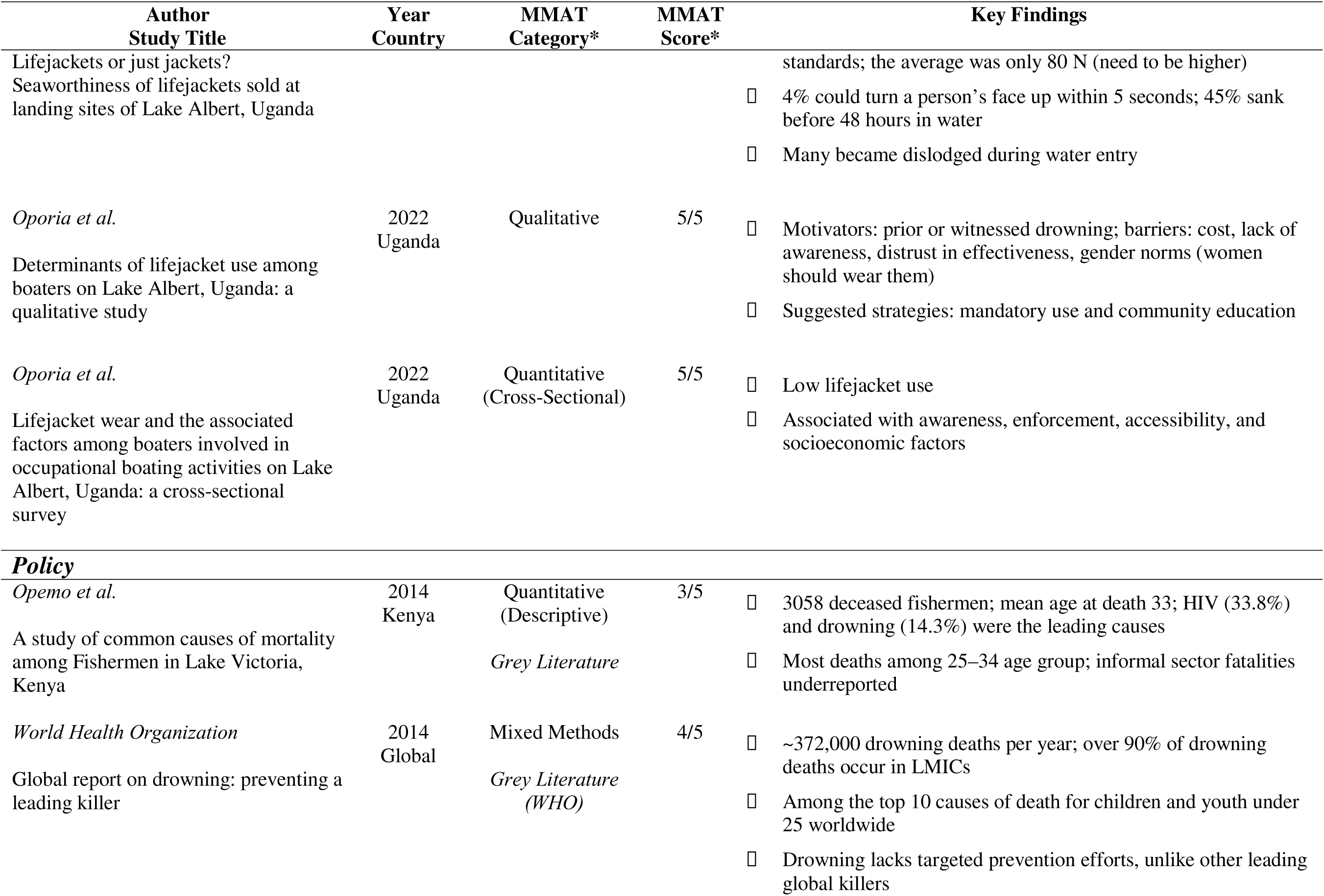
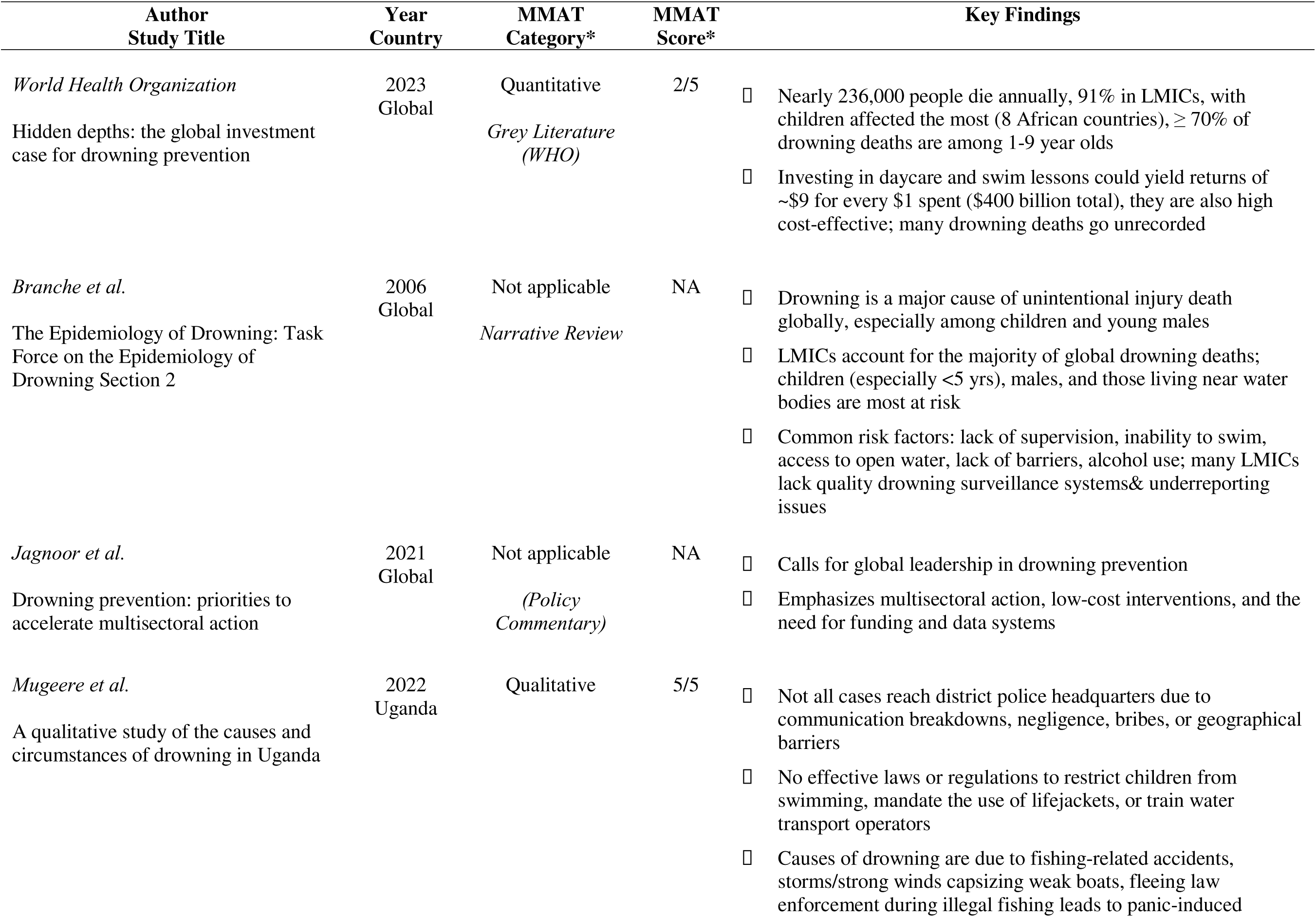
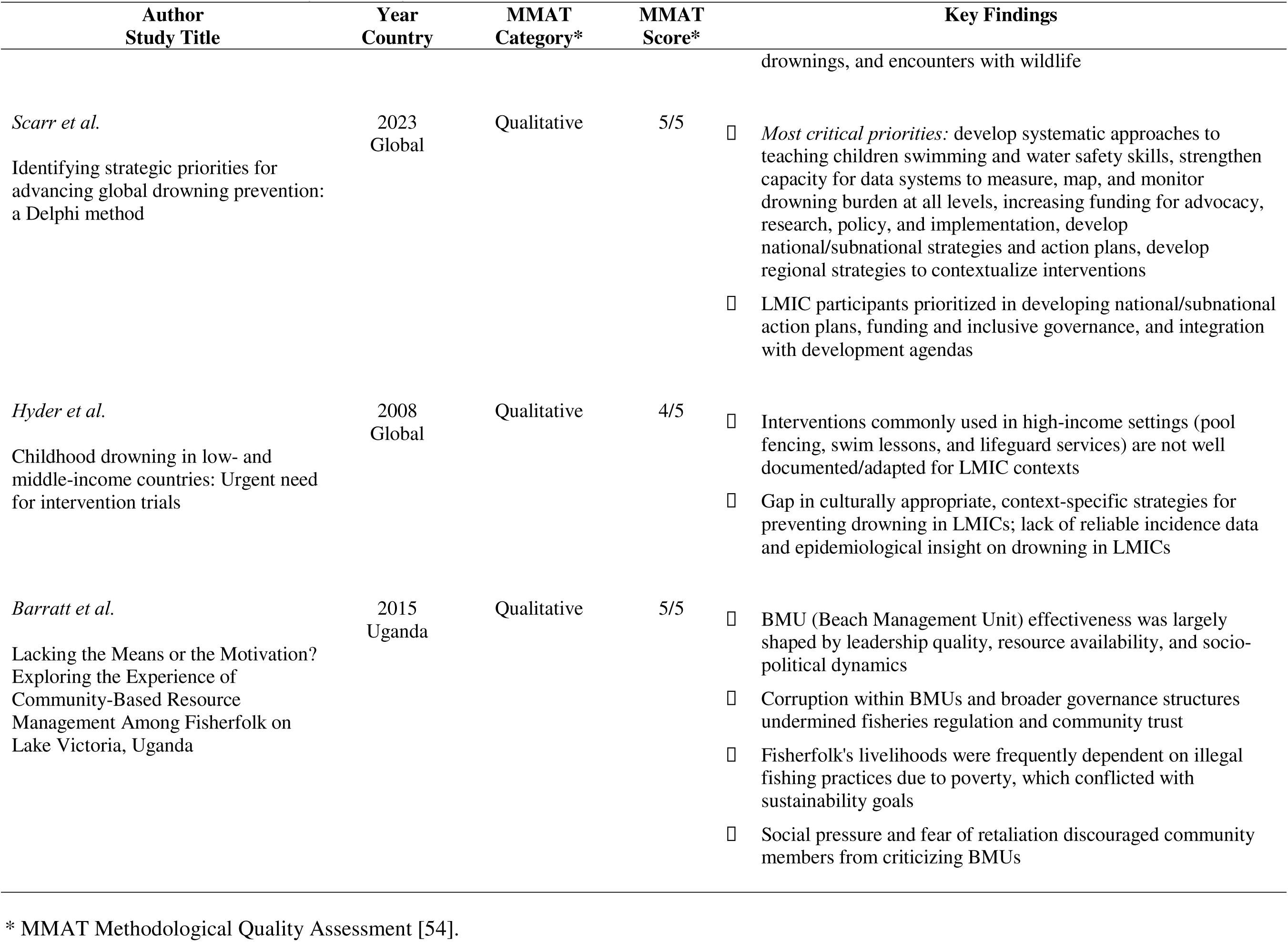
Data Extraction for Study & Quality Assessment.

## Results

Investigators identified 43 studies, which included 37 peer-reviewed articles on drowning in Uganda, Kenya, or Tanzania. Articles were synthesized by year of publication, country setting, geographic scope within a country (if applicable), “prevalence,” “risk factors,” or “policy” tag, and mention of community knowledge of attitudes. Eight studies focused on drowning globally - of these, one focused on risk factors, two on prevalence, and five on policy. Seven studies focused on more than one of the Lake Victoria basin countries, often the result of studies with sites at lakeside communities spanning across multiple countries. Twenty studies were conducted in Uganda alone, three in Kenya, and five in Tanzania. Uganda is the country of focus for 46.5% (n=20) of the research analyzed (Figure 2). Only 17.9% (n=5) of the studies analyzed had a focus on local populations, with regional studies predominating at 67.9% (n=19). Of studies focusing on just one country, 14.3% (n= 4) had a national scope.

**Figure 2:**
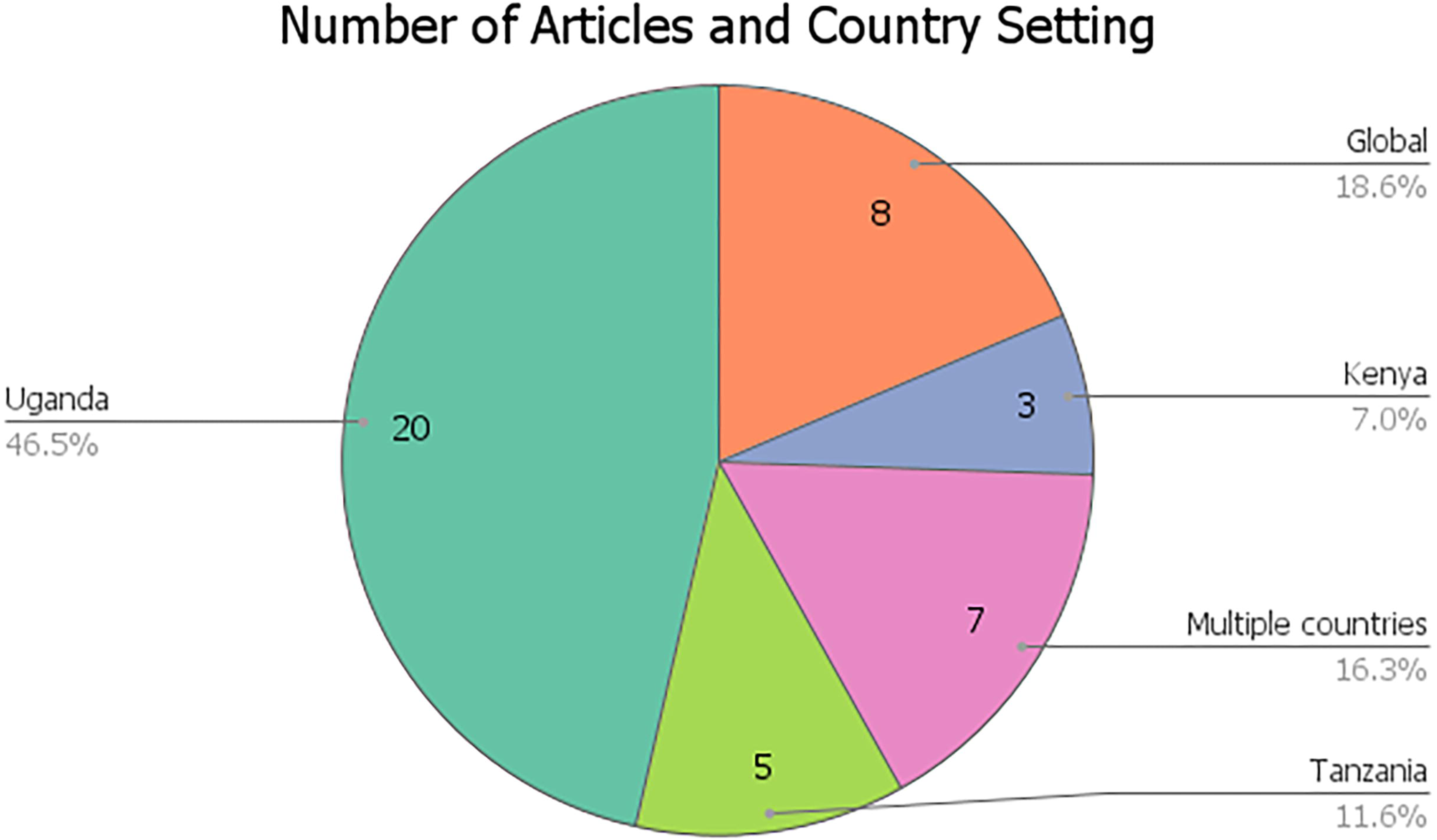
Number of articles and country setting; Multiple countries were used as an indicator for any article focused on some combination of the three countries of focus (Tanzania, Uganda, Kenya).

Twelve of the 37 peer-reviewed articles on drowning used qualitative methodologies such as interviews, focus groups, and non-statistical methods [8, 13–14, 16, 18, 20, 25, 27, 31–32, 39, 46]; ten had quantitative [9, 11, 17, 19, 28, 36–38, 40, 45]. Three were cross-sectional studies [10, 15, 47], and another six were mixed-methods [4, 21–22, 33, 42, 44]. One randomized control trial [43], one narrative review [28], two observational studies [34–35], and two systematic reviews [50, 23] were included in the analysis. Only 30.2% (n=13) of the articles analyzed had any mention of community knowledge, beliefs, or attitudes regarding drowning. When analyzed in terms of content, over half of the research articles examined focused on incidence/prevalence (23.2%, n=10) or risk factors (41.8%, n=18)(Figure 3). Just 6 of the articles studied had a focus on direct interventions– mixed-methods or randomized trials which directly tested a resource, such as lifejackets, or a training, such as water safety training to assess effectiveness directly– with many of these studies conducted on Lake Albert, not Lake Victoria (13.9%, n=6).

**Figure 3:**
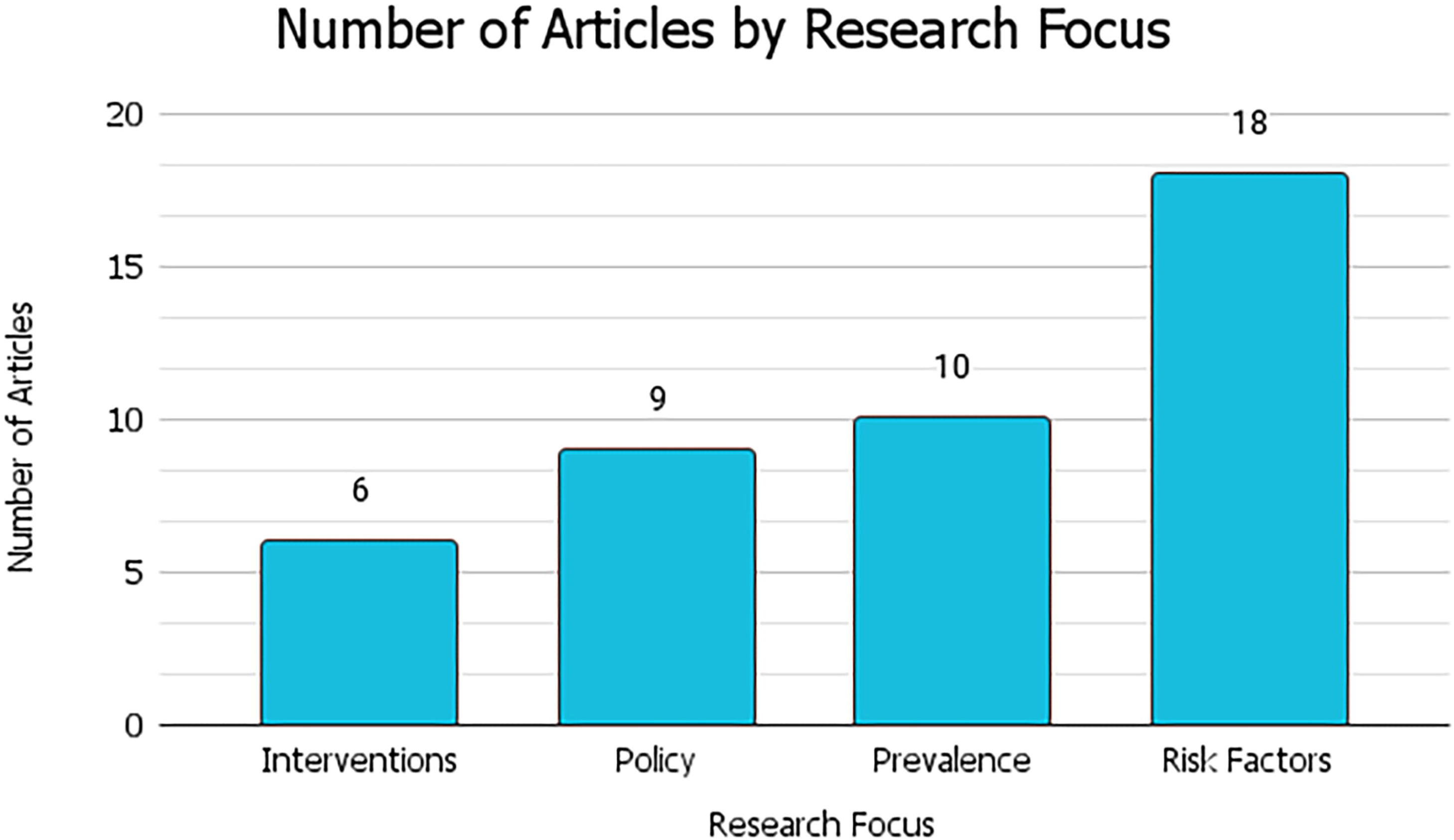
Research focus of articles selected for analysis.

**Figure 4:**
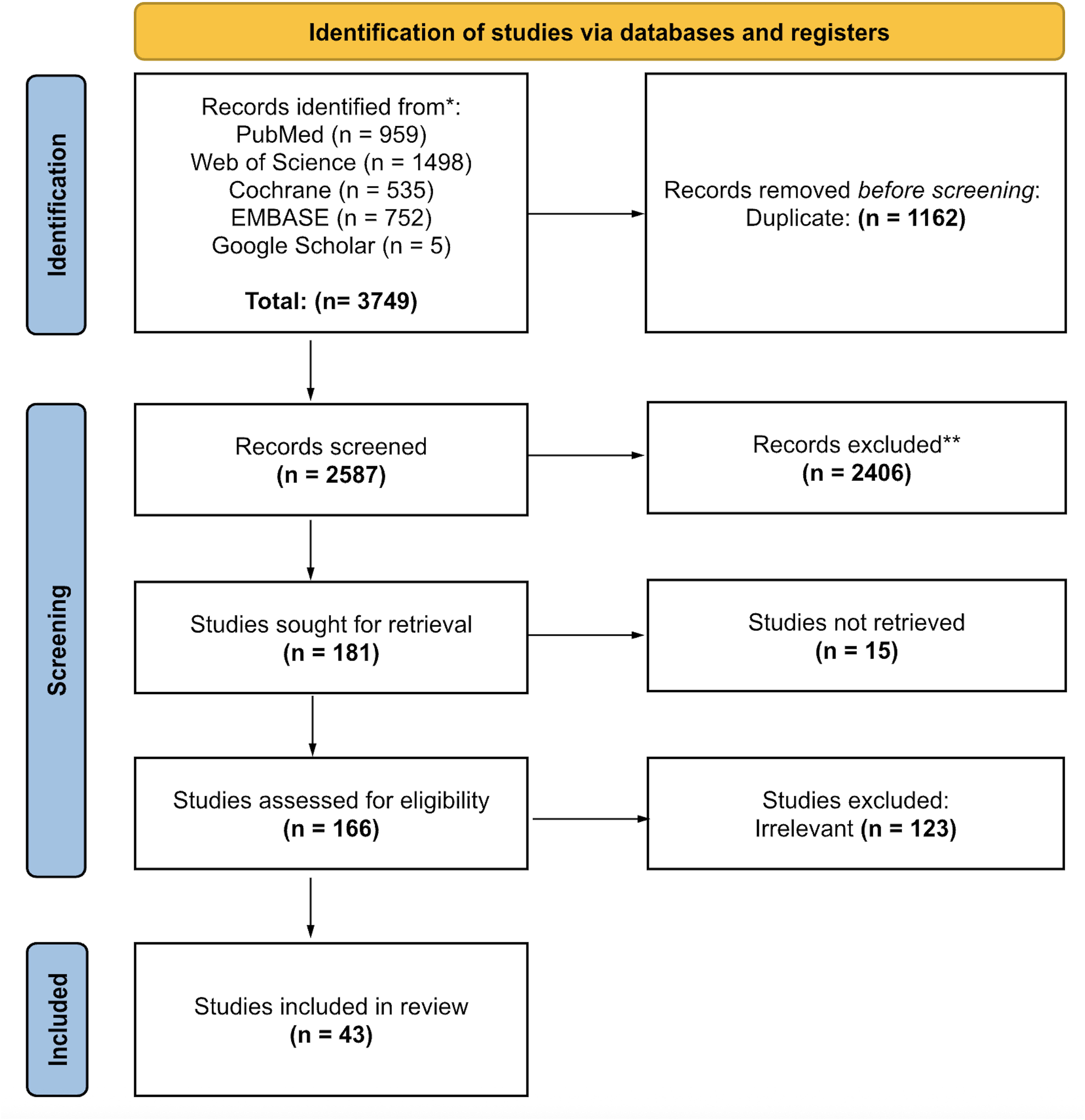
Methodology of articles that were peer-reviewed. Note: Gray literature such as non-profit reports were excluded from this analysis.

### Prevalence

Five studies have reported the prevalence of drowning through various methods such as questionnaires and in-person interviews [33–34], combining population-based data [36], and World Bank and WHO data [39]. One study based in Uganda found a drowning fatality rate of 502 deaths per 100,000 population in the four Uganda lakeside communities [33].

One Tanzanian study using 13 data sources found a prevalence of 5.1 drowning deaths per 100,000 persons per year in Tanzania [36]. Another study in northwest Tanzania estimated the incidence of drowning to be 217/100,000 population [34]. A population-based study in two rural and one urban community in Tanzania found that the prevalence of unintentional drowning injury and death was 17.1 per 100,000 and 6.9 per 100,000, respectively. Another study estimated that there are 2100 to 7700 drowning deaths per year occurring in the Tanzanian lakeside fishing communities alone [34]. In a rural district in Kenya, 17% of all deaths among persons ages 15-64 years in the 1980s were attributed to injuries, though not specifically to drowning [39].

### Risk Factors

Several publications reported on identified risk factors of drowning, those consistently being: age, male gender, alcohol use, household and residence, access to bodies of water, fishing as occupation, boating transportation and carry capacity, boat maintenance, and unpredictable weather patterns.

One article examined gender as a being risk factor for drowning and found males were at greater risk than females for drowning [36]. Two studies in Uganda and two in Tanzania identified young age, less than 10 years, as a significant risk factor for drowning [19, 21, 42, 36]. Conversely, one study in Tanzania identified adults 45 or older as being at greater risk of drowning [36]. Another records examination study in Uganda found young adulthood of about 24 years old was also an age risk factor, perhaps in association with occupational drowning deaths [37]. A study in Tanzanian lakeside fishing communities found the majority of persons dying from drowning were fishermen aged 18 – 40 years who died while fishing at night, most in small wooden boats powered only by paddles [34].

With respect to occupation as a risk factor, two studies in Uganda identified boating as transportation or fishing as a risk factor for drowning [33, 37]. In a study of eight fishing communities on the Tanzanian shoreline of Lake Victoria, over 80% of drowning deaths were among fishermen (all males) [42]. Three studies in Uganda identified rural households or residences as a risk factor for drowning [11, 37–38]. In Tanzania, a population-based study found that drowning was the leading cause of injury and death in rural areas and was the second leading cause of injury and death in males under 5 years old in the city of Dar es Salaam [38]. Multiple studies have linked alcohol use with occupation as a drowning risk factor. In the population-based study in Tanzania, the use of alcohol played a role in fatal drownings, with 64.6% of all adult victims testing positive for blood alcohol content (BAC) [38].

In a survey of 316 fishermen and boat owners and operators in Uganda, Kenya, Tanzania, and Rwanda, as well as 5 anecdotal interviews at the Ggaba landing site near Kampala, Uganda, fishermen cited poor boat maintenance, boat overloading, and weather events as risk factors [13]. Research conducted with 1000 respondents at 8 sites on Lake Victoria in Tanzania cited risk factors of unstable vessels and changeable conditions, along with an inability to cope or call for help [24]. Out of another survey (n = 316) of fishermen, boat owners, and operators in Uganda, Kenya, Tanzania, and Rwanda, only 59.8% of respondents knew where the weather station was located in their region [13]. In a study of 215 respondents interviewed on 14 landing sites on Lake Victoria, 92% (198/215) of the respondents reported using mobile phones as their main tool of communication, but only 4% had smartphones that could receive early warning weather alerts through internet connectivity [10]. Findings from another study’s focus groups in Kenya and Uganda in mid-2020 indicated that 75% of those who receive weather information use it to inform if and when to travel on the lake [9].

### Drowning Prevention Intervention Strategies

Drowning prevention intervention strategies and recommendations studied in the literature included life jacket use and peer-led water safety training. All sources presenting drowning prevention intervention strategies reported improving life jacket usage. Three studies in Lake Albert, Uganda, as well as one study on the Tanzanian shoreline of Lake Victoria, recommended the same interventions: improved life jackets, boat maintenance, swimming lessons, and/or life jacket training [42, 37, 47, 44]. One study recommended having awareness campaigns and lights on boats [42]. Another study suggested following the WHO Global Report on Drowning interventions such as enforcing safe boating practices and implementing more preventive interventions [33].

A two-arm cluster randomized controlled trial across 14 landing sites in Uganda demonstrated that a peer-led training program significantly increased life jacket use among occupational boaters on Lake Albert. After the program, life jacket wear increased from 30.8% at baseline to 65.1%, with 81.5% of boaters reporting they intended to continue wearing life jackets during future trips [43]. In another study of 544 participants from four districts in Uganda, one-third (33.9%) of respondents reported owning a life jacket, and two-thirds (67%) had used a life jacket at some stage [33].

### Current Policy Recommendations

There were few articles with the primary aim of policy analysis or policy investigation. Globally, one study which included research from Tanzania and Uganda developed 10 strategic priorities for advancing drowning prevention, including systematic swimming and water safety education, strengthening data capacity to monitor burden, developing national drowning prevention plans, and strengthening national coordination [27]. These priorities could be used by funders, researchers, and national coalitions to make investment decisions, develop research questions, and develop effective drowning prevention policies. [27].

One regional qualitative study found that policy recommendations in Uganda included the creation of specialized drowning reporting systems, and transitioning from paper-based records to a shareable national network [25].

One local study in Uganda found that despite the increased lakeshore earnings, certain groups benefited more because of local communities competing with international markets [32]. The study reveals disparities in benefits and corruption in the Kitanba Beach Management Units (BMU). Though the BMU model reduced the fishery management burden on the government, lack of equipment, training, as well as the voluntary nature of BMU positions were all noted as factors that impeded the effectiveness of the BMU model. While some BMUs reported improvements in sanitation and reduced illegal fishing, corruption within committees allowed illegal practices to persist due to diminishing fish stocks [32].

## Discussion

This systematic review aimed to provide an overview of literature on drowning in the Lake Victoria basin and associated risk factors, interventions, policy, and prevalence of drowning in this region. A total of 43 studies were identified, with 36 of 43 being peer-reviewed articles and 7 being classified as gray literature. While there has been an increase in publications regarding drowning, significant research gaps exist for the Lake Victoria basin including a lack of studies with a geographic focus in Kenya or Tanzania, studies of local populations, studies of direct interventions against drowning, studies on behavioral (non-demographic) risk factors such as alcohol use, and studies centered on identifying community beliefs and values around drowning.

While the Lake Victoria Basin consists of Uganda, Kenya, and Tanzania, analysis of the literature’s country of focus found limited research in Tanzania and Kenya. Tanzania has the greatest percentage of the Lake Victoria shoreline within its borders (51%) but represents only 11.6% (n=5) of research on drowning in the region, whereas Uganda is the country of focus for 46.5% (n=20) of the research analyzed. Though the rates of drowning in Uganda were found to be higher than in Tanzania, lakeside communities in both of these countries exemplify a disproportionately high drowning rate when compared to both their national and global average rates. Furthermore, only a small number of studies (n=5, 17.9%) focused on local populations, with n<100 or geographic scope limited to (at most) one district or both. This analysis suggests not only a gap in research content but also geography, highlighting the need for more research surrounding lakeside fishing communities in Tanzania and Kenya, as well as more studies focused on local populations.

With respect to prevalence, one study based in Uganda found a drowning fatality rate of 502 deaths per 100,000 population in the four Uganda lakeside communities [33]. This represents a prevalence of drowning deaths that is almost sixty-three times greater than the WHO estimated prevalence of drowning for the WHO Africa region. Another study in northwest Tanzania estimated the incidence of drowning to be 217/100,000 population, 43-fold higher than the estimated annual drowning mortality rate for Tanzania as a whole [34]. Vast discrepancies between prevalence and incidence rates found in the literature compared to drowning prevalence as cited by national averages may reflect an underreporting, misclassification, or lack of consistent surveillance of drowning deaths in the Lake Victoria basin. Discrepancies could exist because of underreporting, misclassification, or lack of consistent surveillance of drowning or rates differ by small regions comparatively with national rates.

Young age, as well as young-adulthood, was frequently identified as being linked with drowning in numerous studies in Uganda and Tanzania. Male gender was also found to be a risk factor in Tanzania [36], suggesting that further research into gender as a demographic risk factor is needed in both Uganda and Kenya and interventions should be developed in accordance with these findings. Similarly, occupation and rural residence were cited as risk factors in research based in both Uganda and Tanzania. Across all the countries in the Lake Victoria basin, boat maintenance and weather were cited as risk factors for drowning, suggesting again the role that occupation plays in drowning risk.

Consistent with these findings, a systematic review of drowning data globally found risk factors of young age (<17-20 years old), male gender (75% vs. 25% female), rural environment (84% vs. 16% urban), occurring in the daytime (95% vs. 5% nighttime), lack of adult supervision (76% vs. 18% supervised), and limited swimming ability (86% vs. 10% with swimming ability). There was an almost equal risk of drowning in a small body of water versus a large body of water (42% ponds, ditches, streams, and wells; 46% lakes, rivers, sea, and ocean) [14, 23].

Occupational boaters and fishermen are at a particularly high risk for drowning based on these risk factors as they are often young adult men, possibly using alcohol, and lacking the ability to swim. In two focus group studies in Uganda, alcohol was cited as a common coping mechanism to deal with work-related stress and both occupational hazards and financial stressors due to inconsistent and unpredictable income dependent on catch yields [18]. Such high rates of alcohol consumption in fishing communities, combined with the preventable lack of water safety knowledge (and the fatalistic perspectives that result from it), have led to a culture where drowning deaths have become accepted as likely or unavoidable [20, 18, 16]. Findings in Tanzania indicate a linkage between alcohol consumption and fatal drownings among fishermen [38]. Occupational boaters and fishermen are also more likely to be in rural areas with fewer water safety resources, including quality boats and education, exacerbating their risk of drowning.

Another significant risk factor presented by the literature was increasing trends of unpredictable weather patterns. In a study of landing site residents in Kalangala, Uganda, 91.7% of respondents (n=215) cited weather, primarily high wind and high waves, as a hazard on the lake [10]. In a study of 8 lakeside communities in Tanzania, of 86 reported drowning deaths in the prior 24 months, 49% occurred during bad weather on the lake [34]. As evidenced by new satellite-based observations and high-resolution climate projections for the African Great Lakes, unpredictable weather patterns on Lake Victoria are projected to continue, emphasizing a need to assess local climate change and activate preventive strategies for fishermen [17]. The use of early weather warning systems was discussed in studies in both Uganda and Kenya. Though they are in place on Lake Victoria, their accessibility and utilization vary by region leaving many communities vulnerable to unpredictable weather. Climate-related warning systems and education must be increased to help fishermen navigate conditions that may continue to worsen.

Current drowning prevention and water safety interventions were analyzed, including behavioral change interventions such as life jacket use. While increasing use of existing life jackets seems to present a low-cost intervention, further research has shown that life jacket quality adds another layer of risk and complexity. A study that tested 22 life jackets from Lake Albert landing sites found that all life jackets failed the minimum buoyancy requirements test to support an adult of at least 40 kg, recommended by local and international standards.

Furthermore, many of the life jackets sold at the landing sites of Lake Albert failed the righting performance test [45]. Of these studies examining direct interventions, three were conducted on Lake Albert, not Lake Victoria. In tandem with a lack of data on local populations, there was a paucity of investigation of community knowledge, beliefs, or attitudes regarding drowning.

Other drowning prevention intervention studies noted that a lack of drowning policy and policy enforcement lends itself to drowning incidence and risk, highlighting the need for comprehensive policy changes. Policy considerations have included those recommended in a Delphi study of strategic priorities for drowning prevention, including restricting alcohol availability around water recreation facilities, separating aquatic incidents from other injuries, never leaving children unattended near water, teaching pool owners CPR, not having telephones near the pool, and having four-sided pool fencing [29]. Multiple studies suggested providing swimming lessons and water safety training for students, as well as recommending children stay away from water through childcare and placing barriers around water hazards [29, 30, 31]. A national plan is needed for interventions to be implemented on a larger scale, and policymakers should ensure flotation devices are available in every village and regulate water safety, boating, and flood disaster risk management [29, 31]. This includes not only the distribution of resources like life jackets, but also community-based interventions to teach swimming, water safety, and proper use of weather warning systems where they are available. Strengthening policy under current knowledge to not only address resource gaps but also the pervasive lack of awareness is crucial for effective drowning prevention.

Other policy considerations include those surrounding the fishing industry. Corruption on Lake Victoria is systemic, involving fishers, police, and judiciary, whom all play a part in the use of illegal equipment, continued overfishing or fishing outside of a country’s territory, and the destabilization of local government’s attempts to regulate, such as Beach Management Units [32, 51]. For the fishing industry, the role of the Ministry of Fisheries in governing and enforcing the International Codes and Guidelines on Occupational Safety and Health is crucial to improving the standards of safety and health in the fishing industry [26].

This review has several strengths and limitations. Given the diverse mixture of included studies, we were unable to provide any summary of the quantitative statistics of prevalence, or risk factors due to large variances in variables and measurement systems. Additionally, relevant studies for Uganda that had a geographic focus on Lake Albert were included, assuming that the results found for that lakeside community are generalizable to the Lake Victoria basin.

Furthermore, this review centers on demographic and behavioral risk factors, but there are many other contributing factors to lakeside drowning as a phenomenon, including socioeconomic determinants as well as medical factors like HIV that contribute greatly to the health landscape of Lake Victoria. No computer-translated foreign-language articles were reviewed for this study - all reviewed articles were available in English. Despite these limitations, to our knowledge, this is the first article that concomitantly reviews drowning policy, interventions, and risk factors for the Lake Victoria basin. This study also includes peer-reviewed literature and a body of non-profit research and dissertation literature that has not been previously summarized, important for developing a profile of drowning incidence and risk data across the Lake Victoria Basin.

### Conclusion

This systematic review revealed several gaps in drowning-related research across the Lake Victoria River Basin Region, including a limited geographic focus in Kenya and Tanzania, available resources, local population dynamics, beliefs about drowning, and behavioral risk factors.

Understanding the existing knowledge and data regarding drowning in the Lake Victoria basin is crucial to planning future research and interventions that will effectively reduce the burden of drowning within lakeside communities. Future research in this region is needed to explore the effectiveness of drowning prevention policies and interventions, evaluate national drowning prevention plans, and examine sociocultural adaptation of interventions. Strengthening surveillance systems and evaluating culturally relevant prevention strategies are critical next steps to address this neglected public health issue. Prioritizing these efforts will support evidence-based interventions aimed at reducing drowning mortality in this high-risk region.

## Declarations

### Registration

The protocol for this systematic review was registered on INPLASY (202560080) and is available in full on inplasy.com (https://doi.org/10.37766/inplasy2025.6.0080).

### Contributorship Statement

MS oversaw design of the systematic review and conducted initial database search in 2/2024 and subsequent title and abstract screening, article analysis, and was a major contributor in writing the manuscript. GG assisted in conducting the updated database search in 6/2025, drafting and updating figures and tables, article analysis, and writing the final manuscript. KG assisted in drafting the final manuscript. FO assisted in developing the search criteria and drafting the final manuscript. HW assisted in the development of the design of the systematic review and assisted with drafting the final manuscript.

### Review Protocol

The protocol for this systematic review was registered on INPLASY (202560080) and is available in full on inplasy.com (https://doi.org/10.37766/inplasy2025.6.0080). PRISMA-P guidelines were followed in the development of this protocol.

### Patient and Public Involvement

Patients or the public WERE NOT involved in the design, or conduct, or reporting, or dissemination plans of our research.

### Competing interests

The authors declare that they have no competing interest / no conflict of interest.

### Ethical Approval

Not applicable as this study was retrospectively analyzing previously collected research.

### Funding

This research received no specific grant from any funding agency in the public, commercial or not-for-profit sectors.

### Data Availability

The authors confirm that the data supporting the findings of this study are available within the article or its supplementary materials.

## Supporting information

Research Checklist

## Acknowledgements

The authors wish to acknowledge Makerere University Trauma, Injury, and Disability (TRIAD) Unit faculty and staff for their contributions and professional insight in the writing of this manuscript. The authors would like to thank the public health students from the University of Southern California for their contributions to the literature review and research dissemination.

## Notes

### Competing Interest Statement

The authors have declared no competing interest.

### Funding Statement

This study did not receive any funding.

