## Supplementary material for "Drowning in the Lake Victoria basin: A systematic review of prevalence, risk factors, and interventions in East Africa": Table 2

| **Table 2.** Data Extraction for Study & Quality Assessment | | | | |
| --- | --- | --- | --- | --- |
| **Author**  **Study Title** | **Year**  **Country** | **MMAT Category*** | **MMAT Score*** | **Key Findings** |
| ***Prevalence*** | | | | |
| *Peden et al.*  World Report on Childhood Injury Prevention | 2008  Global | Not applicable  *Grey Literature*  *(WHO) - narrative* | NA | - Drowning is among the top causes of unintentional childhood death globally, with high rates among 0 – 4 years old & LMICs - Pool fencing, covering hazardous water bodies, wearing personal flotation devices, and CPR can significantly reduce drowning risk - Baby bath seats and solar pool covers may increase the risk of drowning in young children - Early-age swimming lessons, alcohol regulation around water, and clinician-led prevention counselling need more evidence to confirm their effectiveness |
| *Kobusingye et al.*  The Global Burden of Drowning: An African Perspective | 2006  Uganda | Quantitative | 5/5 | - Drowning is significantly underreported due to weak surveillance systems & lack of dedicated injury registries high burden in Africa - Many drownings are preventable, but there is low public awareness |
| *Kobusingye et al.*  Drowning among the lakeside fishing communities in Uganda: results of a community survey | 2016  Uganda | Mixed Methods | 5/5 | - High burden of drowning (502/100,000) - Low life jacket use - Most events during fishing/transport - Overloading and weather cited |
| *Whitworth et al.*  Drowning among fishing communities on the Tanzanian shore of Lake Victoria: a mixed-methods study to examine incidence, risk factors and socioeconomic impact | 2019  Tanzania | Mixed Methods | 4/5 | - High drowning incidence linked to lack of lifejackets, unsafe boats, and alcohol use - Economic burden on families - Recommended regulatory safety interventions |
| *Sarrassat et al.*  Estimating drowning mortality in Tanzania: a systematic review and meta-analysis of existing data sources | 2018  Tanzania | Quantitative | 4/5 | - Males experienced significantly higher drowning rates than female (8.3 vs 3.9/100,000) - Elevated risk among children under 5 & adults over 45 - Average drowning mortality was 5.1/100,000 population per year |
| *Clemens et al.*  Drowning in Uganda: examining data from administrative sources | 2021  Uganda | Quantitative *(Descriptive)* | 3/5 | - No single administrative source in Uganda captured all drowning cases; data completeness varied widely - 70% of identified drowning cases were from lakeside districts, but 30% occurred in non-lakeside districts - Of 1435 cases, 90% were fatal, with males and young children being disproportionately affected |
| *Szpilman et al.*  Drowning prevention, rescue, treatment | 2012  Multiple Countries | Not applicable  *Methodological review/commentary* | NA | - < 6% of all rescued people by a lifeguard need medical attention & 0.5% need CPR - Hospitalization is recommended for patients with a presentation of grade 2 to 6 - A series of hospitalized cases shows that 12% of the people rescued from drowning had pneumonia and needed treatment with antibiotic agents |
| *Forjuoh et al.*  Injury control in africa: Getting governments to do more | 2002  Multiple Countries | Not applicable  *(Commentary Paper)* | NA | - Injury is a leading cause of death and disability in Africa, especially among children & economically active populations - Drowning is under-recognized (limited data collection & public health infrastructure for prevention) - Government focuses on infectious disease over injury prevention - Insufficient research funding, legislation, enforcement, and public awareness regarding injury prevention |
| *Franklin et al.*  The burden of unintentional drowning: global, regional and national estimates of mortality from the Global Burden of Disease 2017 Study | 2020  Global | Quantitative | 4/5 | - Annual deaths fell by 44.5%; age-standardized mortality rate decreased by 57.4% - Mortality rates were highest among children, males, and residents of LMICs; declines in drowning mortality were not consistent globally |
| *Guy et al.*  Mixed-methods community assessment of drowning and water safety knowledge and behaviours on Lake Victoria | 2024  Uganda | Mixed Methods | 5/5 | - Nearly all respondents (93.5%) collect water; 81.4% travel via the lake; 56.4% use it for services; 50% for recreation; only ~30.9% reported at least one family member can swim; 64.2% reported no signage/fences, 95.8% reported no aquatic emergency response system; 85.7% of households experienced a drowning incident in their area - Limited awareness of water safety, participants expressed interest in interventions (swimming lessons, lifejacket use, etc) |
| ***Risk Factors*** | | | | |
| *Pando et al.*  Drowning deaths among fishing communities of Lake Victoria, Tanzania | 2018  Tanzania | Mixed Methods  *Grey Literature* | 5/5 | - 231 drowning deaths per 100,000 (30 times the national/continental average); 81% of deaths were males aged 18-40, 10% children, 9% non-fishing adults, most drowned while fishing - *Risk factors:* poor vessel condition, fishing at night or in bad weather, no lifejacket use (<5%), alcohol/drug use, lack of swimming ability - *Perceived risks and barriers:* low trust in lifejackets, safety not prioritized, stigma around not knowing how to swim - *Socioeconomic impact:* loss of income, displacement, family breakdowns, and community distress |
| *Opemo et al.*  Causes of mortality among the fishermen in Lake Victoria Kenya | 1999  Kenya | Quantitative  *Grey Literature* | 5/5 | - Top causes of death: HIV-related illness (33.8%), drowning (14.3%), pulmonary TB (12.4%), malaria (10.4%); drowning is most frequent among 15-19 year old (27.3% of deaths in this group) - Risk factors include alcohol use, sailing/paddling, and younger age; high proportion of deaths occurred outside health facilities (>90%) |
| *Tushemereirwe et al.*  The Most Effective Methods for Delivering Severe Weather Early Warnings to Fishermen on Lake Victoria | 2017  Uganda | Quantitative  (Cross-Sectional) | 4/5 | - Frequent lake travel and exposure, mobile phones were a trusted source, preference for alarm warnings - 91.7% cited severe weather hazards; 30.7% received alerts, most found helpful; 75% were willing to receive, and 65% to pay - Limitations: convenience sampling and recall bias |
| *Kobusingye et al.*  Injury patterns in urban and rural Uganda | 2001  Uganda | Quantitative | 5/5 | - Urban areas had a significantly higher injury mortality (217/100,000/yr) & disability prevalence (2.8%) than rural areas (92/100,000/yr); the leading cause of death (rural): drowning - Most non-fatal injuries occurred at home or on the roads |
| *Kiwanuka-Tondo et al.*  Climate risk communication of navigation safety and climate conditions over Lake Victoria basin: Exploring perceptions and knowledge of indigenous communities. | 2019  Uganda | Qualitative | 4/5 | - Limited risk communication and preparedness for adverse weather - Fishers’ knowledge gaps hinder risk response - Recommendations include participatory risk communication strategies, culturally tailored warning systems, and integration of indigenous knowledge in climate adaptation |
| *Wandera et al.*  Alcohol use, intimate partner violence, and HIV sexual risk behavior among young people in fishing communities of Lake Victoria, Uganda | 2021  Uganda | Quantitative (Cross-Sectional) | 4/5 | - Alcohol use is linked to increased IPV and HIV sexual risk behavior - Young men in fishing communities face elevated health risks |
| *Kuteesa et al.*  ‘We shall drink until Lake Victoria dries up’: Drivers of heavy drinking and illicit drug use among young Ugandans in fishing communities | 2021  Uganda | Qualitative | 5/5 | - Substance use is widespread among youth in Ugandan fishing communities, especially heavy drinking and marijuana use - Economic insecurity drives alcohol and drug use as a coping mechanism because fishing income is unpredictable - Peer influence and social norms strongly reinforce substance use, with drinking and smoking being seen as an adult identity and masculinity - Lack of regulation enforcement on alcohol and drug sales is making it easy to access - Psychosocial stressors such as exposure to violence, trauma, and family breakdown also contribute to substance use |
| *Thiery et al.*  Hazardous thunderstorm intensification over Lake Victoria | 2016  Multiple Countries | Not applicable  *Climate Modelling & Remote Sensing Study* | NA | - Satellite observations and regional climate model simulations reveal that Lake Victoria induces powerful nocturnal thunderstorms - Lake’s unique land-lake breeze dynamics amplify convective storms, particularly at night - Intensified storms are responsible for frequent lightning, high winds, and extreme rainfall, posing risks to artisanal fishers |
| *Sileo et al.*  “Such Behaviors Are Not in My Home Village, I Got Them Here”: A Qualitative Study of the Influence of Contextual Factors on Alcohol and HIV Risk Behaviors in a Fishing Community on Lake Victoria, Uganda | 2016  Uganda | Qualitative | 5/5 | - Heavy alcohol use and commercial sex work are prevalent and economically embedded in the fishing community - Alcohol is commonly used during sex among CSWs and clients, contributing to risky sexual behavior and limiting condom negotiation - Lack of savings mechanisms, poverty, and weak policy enforcement exacerbate risk; Interventions must target economic, structural, and individual factors |
| *Batte et al.*  Incidence, patterns and risk factors for injuries among Ugandan children | 2017  Uganda | Quantitative  *(Cross-sectional)* | 5/5 | - 113 injuries reported in past 6 months (9.4%); leading causes of injury: falls (43.4%), burns (17.7%), cuts (13.3%), road traffic injuries (8.9%) - Risk factors: 5-9 yrs had the highest injury risk (aOR: 2.21), male children & those from large households are more likely to be injured |
| *Breur et al.*  The Bottle Is My Wife: “Exploring Reasons Why Men Drink in Ugandan Fishing Communities” | 2019  Uganda | Qualitative | 5/5 | - Alcohol use was linked to gender roles, coping mechanisms, and social pressure - Structural and cultural factors contributed to high alcohol consumption. |
| *Kananura et al.*  Under 10 mortality patterns, risk factors, and mechanisms in low resource settings of Eastern Uganda: An analysis of event history demographic and verbal social autopsy data | 2020  Uganda | Mixed Methods | 5/5 | - Leading causes of death include malaria, birth complications, and drowning - Social autopsy revealed care-seeking barriers |
| *Tyler et al.*  The epidemiology of drowning in low-and middle-income countries: a systematic review | 2017  Multiple LMICs | Not applicable  *(Systematic Review)* | NA | - Drowning is a leading cause of death in LMICs - Risk factors include age, gender, and poor water safety - Surveillance and interventions vary widely |
| *Miller et al.*  Epidemiology, Risk Factors and Measures for Preventing Drowning in Africa: A Systematic Review | 2019  Multiple LMICs | Not applicable  *(Systematic Review)* | NA | - Drowning burden in Africa is high; few intervention evaluations exist - Data gaps and the need for stronger prevention strategies were noted |
| *Powell et al.*  Understanding the Risks to Artisanal Fishers on Lake Victoria Using Design Methods | 2025  Tanzania | Mixed Methods | 5/5 | - High drowning risk for both long-line (day) and seine net (night) artisanal fishers - Visual journey risk maps identified hazards before, during, and after fishing trips - Common risks include poor weather prediction, lack of lifejacket use, boat collisions, fatigue, and equipment failure |
| *Roberts et al.*  Taking the HIGHWAY to Save Lives on Lake Victoria | 2022  Multiple Countries | Mixed Methods | 5/5 | - Fishers and transporters lack timely, localized, and actionable weather info - Users expressed concerns about phone charging, network coverage, and trust in forecasts - Enhanced early warning systems are feasible and well-received, but sustained impact will require infrastructure, training, and policy support |
| *Rasolofoson et al.*  Climate change: A pointer to increased small-scale fisher drowning deaths | 2024  Kenya | Mixed Methods | 4/5 | - Bad weather contributed to 41.8% of fatal drownings among small-scale fishers on Lake Victoria - During bad weather incidents, 69.5% of drownings involved no lifejacket, 67.8% lacked navigation equipment - ~57% of deceased fishers could swim; alcohol was present in 31%, drug use in 24% of cases; most victims were men in their 30s, experience fishers with over 3 yrs of experience |
| *Kiwanuka-Tondo et al.*  An assessment of fishing communities around Lake Victoria, Uganda, as potential populations for future HIV vaccine efficacy studies: an observational cohort study | 2019  Uganda | Quantitative (Cohort) | 5/5 | - HIV incidence is high (3.39/100 PYAR) - Retention rate of 77%, high willingness to join trials - Willingness to participate in HIV vaccine trials was higher in men than women (91.2% vs 87.3%, p = 0.004) |
| ***Interventions*** | | | | |
| *Pando et al.*  An observational study to inform potential drowning intervention strategies among fishing communities in the lake zone of Tanzania (DRIFT) | 2018  Tanzania | Mixed Methods | 5/5 | - Identified fishing-related risk behaviors and barriers to safe practices - Emphasized the role of community-driven surveillance, local data collection systems, and the need for stakeholder-informed intervention designs |
| *Oporia et al.*  Peer-led training improves lifejacket wear among occupational boaters: Evidence from a cluster randomized controlled trial on Lake Albert, Uganda | 2023  Uganda | Quantitative | 5/5 | - Self-reported lifejacket uses increased from 30.8% to 65.1% in the intervention group compared to 29.9% to 43.2% in the control group - Directly observed lifejacket use increased from 1.0% to 26.8% in the intervention group vs. control group (0.6% to 8.8%) - After adjusting for contamination, recipients of peer-led training were 1.78 times more likely to report wearing a lifejacket (95% CI 1.38-2.30) |
| *Oporia et al.*  Development and validation of an intervention package to improve lifejacket wear for drowning prevention among occupational boaters on Lake Albert, Uganda | 2023  Uganda | Mixed Methods | 4/5 | - Pilot-tested a co-created intervention improving knowledge, attitudes, and lifejacket use - Culturally tailored interventions were found to be feasible and effective |
| *Oporia et al.*  Lifejackets or just jackets? Seaworthiness of lifejackets sold at landing sites of Lake Albert, Uganda | 2024  Uganda | Quantitative | 4/5 | - 22 tested lifejackets failed to meet minimum buoyancy standards; the average was only 80 N (need to be higher) - 4% could turn a person’s face up within 5 seconds; 45% sank before 48 hours in water - Many became dislodged during water entry |
| *Oporia et al.*  Determinants of lifejacket use among boaters on Lake Albert, Uganda: a qualitative study | 2022  Uganda | Qualitative | 5/5 | - Motivators: prior or witnessed drowning; barriers: cost, lack of awareness, distrust in effectiveness, gender norms (women should wear them) - Suggested strategies: mandatory use and community education |
| *Oporia et al.*  Lifejacket wear and the associated factors among boaters involved in occupational boating activities on Lake Albert, Uganda: a cross-sectional survey | 2022  Uganda | Quantitative (Cross-Sectional) | 5/5 | - Low lifejacket use - Associated with awareness, enforcement, accessibility, and socioeconomic factors |
| ***Policy*** | | | | |
| *Opemo et al.*  A study of common causes of mortality among Fishermen in Lake Victoria, Kenya | 2014  Kenya | Quantitative (Descriptive)  *Grey Literature* | 3/5 | - 3058 deceased fishermen; mean age at death 33; HIV (33.8%) and drowning (14.3%) were the leading causes - Most deaths among 25–34 age group; informal sector fatalities underreported |
| *World Health Organization*  Global report on drowning: preventing a leading killer | 2014  Global | Mixed Methods  *Grey Literature (WHO)* | 4/5 | - ~372,000 drowning deaths per year; over 90% of drowning deaths occur in LMICs - Among the top 10 causes of death for children and youth under 25 worldwide - Drowning lacks targeted prevention efforts, unlike other leading global killers |
| *World Health Organization*  Hidden depths: the global investment case for drowning prevention | 2023  Global | Quantitative  *Grey Literature (WHO)* | 2/5 | - Nearly 236,000 people die annually, 91% in LMICs, with children affected the most (8 African countries), ≥ 70% of drowning deaths are among 1-9 year olds - Investing in daycare and swim lessons could yield returns of ~$9 for every $1 spent ($400 billion total), they are also high cost-effective; many drowning deaths go unrecorded |
| *Branche et al.*  The Epidemiology of Drowning: Task Force on the Epidemiology of Drowning Section 2 | 2006  Global | Not applicable  *Narrative Review* | NA | - Drowning is a major cause of unintentional injury death globally, especially among children and young males - LMICs account for the majority of global drowning deaths; children (especially <5 yrs), males, and those living near water bodies are most at risk - Common risk factors: lack of supervision, inability to swim, access to open water, lack of barriers, alcohol use; many LMICs lack quality drowning surveillance systems& underreporting issues |
| *Jagnoor et al.*  Drowning prevention: priorities to accelerate multisectoral action | 2021  Global | Not applicable  *(Policy Commentary)* | NA | - Calls for global leadership in drowning prevention - Emphasizes multisectoral action, low-cost interventions, and the need for funding and data systems |
| *Mugeere et al.*  A qualitative study of the causes and circumstances of drowning in Uganda | 2022  Uganda | Qualitative | 5/5 | - Not all cases reach district police headquarters due to communication breakdowns, negligence, bribes, or geographical barriers - No effective laws or regulations to restrict children from swimming, mandate the use of lifejackets, or train water transport operators - Causes of drowning are due to fishing-related accidents, storms/strong winds capsizing weak boats, fleeing law enforcement during illegal fishing leads to panic-induced drownings, and encounters with wildlife |
| *Scarr et al.*  Identifying strategic priorities for advancing global drowning prevention: a Delphi method | 2023  Global | Qualitative | 5/5 | - *Most critical priorities:* develop systematic approaches to teaching children swimming and water safety skills, strengthen capacity for data systems to measure, map, and monitor drowning burden at all levels, increasing funding for advocacy, research, policy, and implementation, develop national/subnational strategies and action plans, develop regional strategies to contextualize interventions - LMIC participants prioritized in developing national/subnational action plans, funding and inclusive governance, and integration with development agendas |
| *Hyder et al.*  Childhood drowning in low- and middle-income countries: Urgent need for intervention trials | 2008  Global | Qualitative | 4/5 | - Interventions commonly used in high-income settings (pool fencing, swim lessons, and lifeguard services) are not well documented/adapted for LMIC contexts - Gap in culturally appropriate, context-specific strategies for preventing drowning in LMICs; lack of reliable incidence data and epidemiological insight on drowning in LMICs |
| *Barratt et al.*  Lacking the Means or the Motivation? Exploring the Experience of Community-Based Resource Management Among Fisherfolk on Lake Victoria, Uganda | 2015  Uganda | Qualitative | 5/5 | - BMU (Beach Management Unit) effectiveness was largely shaped by leadership quality, resource availability, and socio-political dynamics - Corruption within BMUs and broader governance structures undermined fisheries regulation and community trust - Fisherfolk's livelihoods were frequently dependent on illegal fishing practices due to poverty, which conflicted with sustainability goals - Social pressure and fear of retaliation discouraged community members from criticizing BMUs |

* MMAT Methodological Quality Assessment [54].
