## Supplementary material for "Drowning in the Lake Victoria basin: A systematic review of prevalence, risk factors, and interventions in East Africa": Table 1

Table 1: Search terms

Search phrase

fishing communities, Lake Victoria, HIV

demographics, Lake Victoria, Uganda

drowning, Lake Victoria, Tanzania

weather, Lake Victoria

world report, drowning

rural Uganda, injury

injury, Uganda, drowning

fishermen, mortality, Kenya

drowning, Tanzania

climate risk, Lake Victoria, accidents

child injury prevention

Kenya, fishing community

Tanzania, drowning interventions, fishing communities

drowning prevention, global drowning prevention plans, risk reduction

epidemiology of drowning, risk factors, burden of drowning, strategies

training, occupational boaters, Uganda

fishing communities, Lake Victoria

fishermen, Lake Victoria

drowning prevention, Africa

drowning, Lake Victoria

drowning

drowning, lake fishing communities

drowning prevention
